# Microstructure-Informed Myelin Mapping (MIMM) from Gradient Echo MRI using Stochastic Matching Pursuit

**DOI:** 10.1101/2023.09.22.23295993

**Authors:** Mert Şişman, Thanh D. Nguyen, Alexandra G. Roberts, Dominick J. Romano, Alexey V. Dimov, Ilhami Kovanlikaya, Pascal Spincemaille, Yi Wang

## Abstract

Quantification of the myelin content of the white matter is important for studying demyelination in neurodegenerative diseases such as Multiple Sclerosis (MS), particularly for longitudinal monitoring. A novel noninvasive MRI method, called Microstructure-Informed Myelin Mapping (MIMM), is developed to quantify the myelin volume fraction (MVF) by utilizing a multi gradient echo sequence (mGRE) and a detailed biophysical model of tissue microstructure. Myelin is modeled as anisotropic negative susceptibility source based on the Hollow Cylindrical Fiber Model (HCFM), and iron as isotropic positive susceptibility source in the extracellular region. Voxels with a range of biophysical parameters are simulated to create a dictionary of MR echo time magnitude signals and total susceptibility values. MRI signals measured using a mGRE sequence are then matched voxel-by-voxel to the created dictionary to obtain the spatial distributions of myelin and iron. Three different MIMM versions are presented to deal with the fiber orientation dependent susceptibility effects of the myelin sheaths: a basic variation, which assumes fiber orientation is an unknown to fit, two orientation informed variations, which assume the fiber orientation distribution is available either from a separate diffusion tensor imaging (DTI) acquisition or from a DTI atlas based fiber orientation map. While all showed a significant linear correlation with the reference method based on T2-relaxometry (p < 0.0001), DTI orientation informed and atlas orientation informed variations reduced overestimation at white matter tracts compared to the basic variation. Finally, the implications and usefulness of attaining an additional iron susceptibility distribution map are discussed.

**Highlights:**

- novel stochastic matching pursuit algorithm called microstructure-informed myelin mapping (MIMM) is developed to quantify Myelin Volume Fraction (MVF) using Magnetic Resonance Imaging (MRI) and microstructural modeling.
- utilizes a detailed biophysical model to capture the susceptibility effects on both magnitude and phase to quantify myelin and iron.
- matter fiber orientation effects are considered for the improved MVF quantification in the major fiber tracts.
- acquired myelin and iron maps may be utilized to monitor longitudinal disease progress.

## 1. Introduction

The myelin sheath is a multilamellar structure of lipid bilayers formed by glial cells around the nerve fibers, which allows rapid electrical conduction between neurons and provides trophic support to the axons (Bean, 2007; Brady, Siegel, Albers, R.W., & Price, 2005). Myelination is an essential process in the development and maturation of the central nervous system, and loss of myelin (demyelination) has been implicated in normal aging as well as in neuroinflammatory and neurodegenerative diseases such as multiple sclerosis (MS) (Dobson & Giovannoni, 2019). Therefore, the development of a non-invasive myelin-specific quantitative biomarker is highly important for the diagnosis, monitoring, and therapeutic management of these diseases.

Myelin water fraction (MWF) mapping is a magnetic resonance imaging (MRI) method that aims to quantify the ratio of the myelin water to the total water content of each voxel (Lazari & Lipp, 2021; J. Lee et al., 2021; A. MacKay et al., 2006; Mackay et al., 1994; A. L. MacKay & Laule, 2016; Mancini et al., 2020; van der Weijden et al., 2021). Traditionally, MWF has been measured by applying multi-component T2-relaxometry to the multi-echo data acquired with a slow 2D multi-echo spin echo (MESE) sequence. More efficient 3D acquisition schemes including the gradient and spin echo (GRASE) (Prasloski et al., 2012) and fast acquisition with spiral trajectory and adiabatic T2prep (FAST-T2) (Nguyen et al., 2016; Nguyen, Spincemaille, Gauthier, & Wang, 2017; Nguyen et al., 2012) have been proposed to reduce the acquisition time for clinical translation. Due to the ill-posed nature of the inverse Laplace transform, these approaches are highly sensitive to noise, which necessitates acquisitions with a large voxel size to improve SNR. Furthermore, MWF measurements obtained by T2-based methods can be confounded by fiber orientations and the presence of iron (Bartels et al., 2022; Birkl et al., 2019; Birkl et al., 2020; Birkl, Doucette, Fan, Hernández-Torres, & Rauscher, 2021).

More recently, there has been a great interest in myelin mapping using the multi-echo gradient echo (mGRE) sequence as it is widely available on clinical scanners and can provide whole-brain isotropic coverage in a short acquisition time with much lower specific absorption rate than spin echo sequences (Haacke, Xu, Cheng, & Reichenbach, 2004; Markl & Leupold, 2012). MWF can be calculated by performing three-compartment T2*-relaxometry of the magnitude (Du et al., 2007; Hwang, Kim, & Du, 2010) or the 3 pool complex fitting (3PCF) (H. Lee et al., 2018; Nam, Lee, Hwang, & Kim, 2015) of mGRE signal, with the latter approach considered more sensitive by utilizing the frequency offsets created by the white matter fiber bundles (Sukstanskii & Yablonskiy, 2014; Wharton & Bowtell, 2012). This approach showed that the mGRE phase is an additional source of information regarding tissue myelin content. Nevertheless, the 3PCF approach requires the non-linear iterative fitting of 10 parameters per voxel, which is slow and sensitive to the choice of parameter initialization and stopping criteria (Chan & Marques, 2020; Nam et al., 2015). Furthermore, this approach overestimates the myelin content in iron-rich brain regions such as basal ganglia since iron also causes rapid signal decay and frequency shifts (Dong et al., 2021; Hédouin et al., 2021; Jung, Yun, Kim, & Kim, 2022).

To improve the performance of mGRE-based MWF imaging, Chan and Marques (Chan, Chamberland, & Marques, 2023; Chan & Marques, 2020) proposed two algorithms that incorporate either diffusion or multi flip angle mGRE measurements on top of the regular mGRE acquisition. Although these algorithms improve myelin quantification, they require long scan times (> 20 min). Recently, Hédouin et al. (Hédouin et al., 2021) proposed to generate a dictionary of complex signal evolutions based on an HCFM simulation of the WM fibers (Wharton & Bowtell, 2012, 2013), which was then utilized to train a neural network for fast parameter extraction. However, this method does not model heterogeneous background susceptibility due to iron and requires multiple orientations in data acquisition that is not feasible in clinical practice.

In this study, inspired by the dictionary generation strategy in (Hédouin et al., 2021) and matching pursuit approaches (Bergeaud & Mallat, 1995; Chen, 2011; Mallat & Zhang, 1992; Tropp & Gilbert, 2007), we propose a novel myelin quantification method to estimate the MVF from a single 6-min mGRE scan (Sisman, Nguyen, et al., 2023; Sisman, Romano, et al., 2023). The central element of this approach is a detailed signal model which connects the macroscopic mGRE signals (T2* magnitude decay and magnetic susceptibility derived from quantitative susceptibility mapping (QSM) (Azuma et al., 2016; A.V. Dimov et al., 2018; T. Liu et al., 2010; Wang & Liu, 2015)) measured on the voxel level to the micro-scale myelin network and iron distribution. Further, we propose a stochastic matching pursuit algorithm for fast and effective dictionary matching. We performed in vivo evaluation of the developed MIMM method by comparing it with the 3PCF algorithm and also with MWF measurements obtained by the FAST-T2 method in healthy volunteers.

## 2. Theory

### 2.1. MIMM Dictionary Generation

Similar to previous approaches, the hollow cylindrical geometries are initially assumed to extend to infinity such that the field perturbations created by these cylinders do not change along their main direction. However, in MIMM, the computed fields are only considered in a 3D grid with fixed orientations, obtained by resampling the replicated 2D fields to the laboratory frame of reference. The 2D field perturbations produced by the hollow cylinders are (Wharton & Bowtell, 2012):

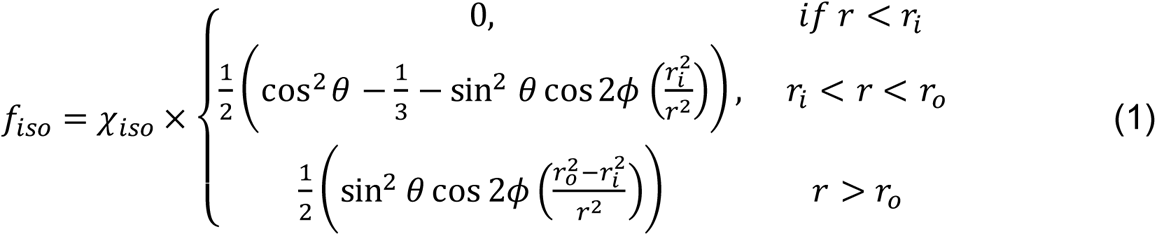

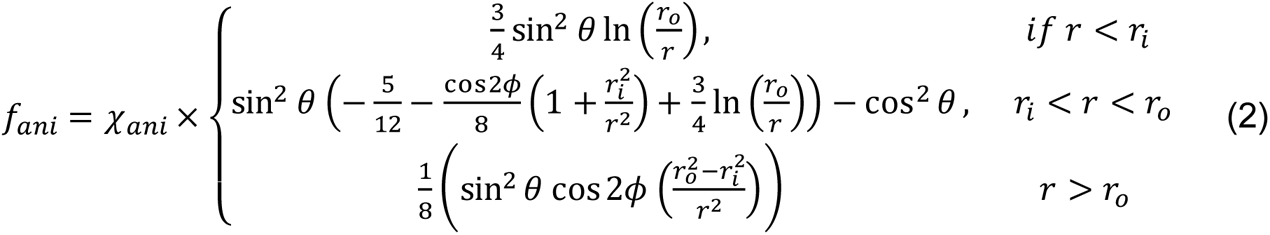

Here, *χ*_*iso*_ and *χ*_*ani*_ are isotropic and anisotropic volume susceptibilities of the myelin sheath, respectively. *f*_*iso*_and *f*_*ani*_ are the field perturbations corresponding to these two susceptibility components. *θ* is the angle between the principal fiber axis and the main magnetic field direction of the MR scanner. *r*_*i*_, and *r*_*o*_ are the inner and outer radii of fibers excluding and including myelin sheath, respectively. *ϕ* and *r* are the cylindrical coordinates in the fiber frame of reference.

In the classical mGRE based MWI approaches, an overestimated level of myelin water in basal ganglia is observed as a systematic artifact (Hédouin et al., 2021; Jung et al., 2022) mainly because of the high iron concentration in these regions. In order to tackle this problem, randomly distributed iron inclusions in the extracellular region are also included in the simulated volume as a second major susceptibility source. Considering that brain iron mainly exists in ferritin form (Wang & Liu, 2015) which is much smaller (∼8 − 12 *nm*) (Clemente-León et al., 2006) than the WM fibers (∼1 *μm*) (Graf von Keyserlingk & Schramm, 1984), iron inclusions are represented by single sub-voxels in the extracellular space. The iron-based field perturbation (*f*_*iron*_) is computed by convolution with the dipole kernel using Fourier transform for computational efficiency. In order to avoid wrapping artifacts, the simulated volume is padded with zeros in each direction before convolution, and the padded regions are cropped afterward. Consequently, the total field perturbation in sub-voxel *j* is:

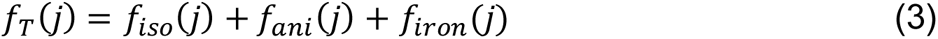

The complex signals corresponding to each sub-voxel for each echo (TE) time are computed and the total signal coming from the whole volume at each TE is computed as:

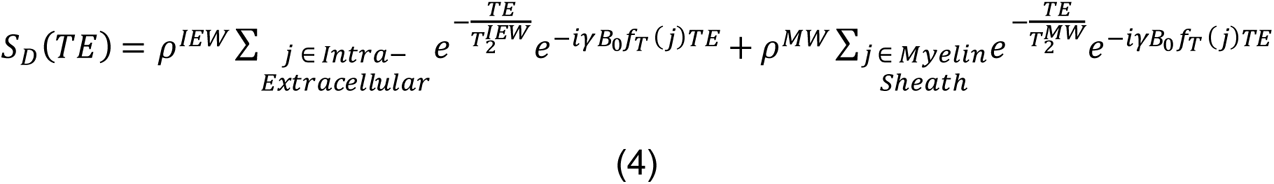

with 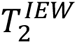, and 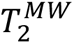 are the transversal relaxation time constants of the intra-extracellular and myelin water compartments, respectively. Similarly, *ρ*^*IEW*^and *ρ*^*MW*^ are the relative proton density of the intra-extracellular and myelin water compartments. *γ* is the gyromagnetic ratio of water proton and *B*_0_is the strength of the static magnetic field of the MR scanner. While some parameters are varied in certain ranges to create the dictionary, some of the parameters are kept constant to decrease the dictionary size and computation time since both increase exponentially with the number of variable parameters. Moreover, it also makes the matching pursuit process more robust considering the fact that fitting for coupled parameters (such as volume susceptibilities and volume fractions) can be a more ill-posed inverse problem. The varying and constant parameters and the corresponding ranges are given in Table 1. The dictionary signal *S*_*D*_was computed for a TE range of [0, 60] *ms* with 3 *ms* step size (21 echoes). In summary, 4 varying input parameters are chosen as:

(1) 2D Fiber Density (2DFD): The ratio of the hollow circles’ area to the whole 2D grid area.
(2) g-ratio: The geometric ratio of the inner radius of the fiber axon to the outer radius (*r*_*i*_/*r*_*o*_).
(3) Extracellular Iron Density (EID): The ratio of the extracellular sub-voxels that are iron inclusions to all extracellular sub-voxels.
(4) Fiber Orientation (*θ*).

**Table 1:** 3D microstructure informed dictionary simulation parameters and the utilized ranges of the parameters. The parameters are shown in 4 different categories based on their functions. 4 input parameters are the parameters that need to be defined for the simulations required to create a dictionary element. Intermediate parameters are the volume fractions that are estimated from the created volume distribution of myelin and iron and utilized to estimate voxel susceptibilities. Output parameters are the labels of each dictionary element that are assigned to each voxel after MIMM. The final set is the constant parameters that have been used in the dictionary generation. The values of the constant parameters are determined using the reported literature values. The corresponding studies for each constant parameter are also provided.

|  | Parameter | Range |
| --- | --- | --- |
| **Dictionary Generation Process Input Parameters** | **2DFD** | [0,1] |
|  | **g – ratio** | [0.5,1] |
|  | **$\theta$** | [0°, 90°] |
|  | **EID** | [0,1] |
| **Intermediate Parameters** | **FVF** | [0, 0.75] |
|  | **IVF** | [0,1] |
| **MIMM Output Parameters** | **MVF** | [0, 0.55] |
|  | **$\chi^{\text{iron}}$** | [0 ppb, 300 ppb] |
| **Constant Parameters** | **$\chi_{\text{iso}} = \chi_{\text{ani}}$** | –100 ppb (Wharton & Bowtell, 2012) |
|  | **$\chi_{\text{iron}}$** | 300 ppb (Zheng, Nichol, Liu, Cheng, & Haacke, 2013) |
|  | **$T_2^{\text{IEW}}$** | 70 ms (S. H. Kolind, Mädler, Fischer, Li, & MacKay, 2009) |
|  | **$T_2^{\text{MW}}$** | 16 ms (Hédouin et al., 2021; Xu, Foxley, Kleinnijenhuis, Chen, & Miller, 2018) |
|  | **$\rho^{\text{IEW}}$** | 1 (Hédouin et al., 2021; Xu et al., 2018) |
|  | **$\rho^{\text{MW}}$** | 0.5 (Hédouin et al., 2021; Xu et al., 2018) |

The steps of the simulation of one dictionary element are summarized in Figure 1a. First, a random distribution of 2D fibers in a circular region is created with varying radii but a fixed g-ratio (Danz, 2020). Then, the field perturbations of each myelin source are computed using Equations 1 and 2 at each sub-voxel. The computed field distribution is extended to a 3D grid by simple replication and resampling based on the fiber orientation. A smaller region of interest (ROI) at the center of the 3D grid is extracted to imitate an imaging voxel where susceptibility sources exist both inside and outside. The fiber volume fraction (FVF) and iron volume fraction (IVF) of the simulated volume are computed as the relative volumes covered by the fibers and iron inclusions within this ROI.

**Figure 1.**
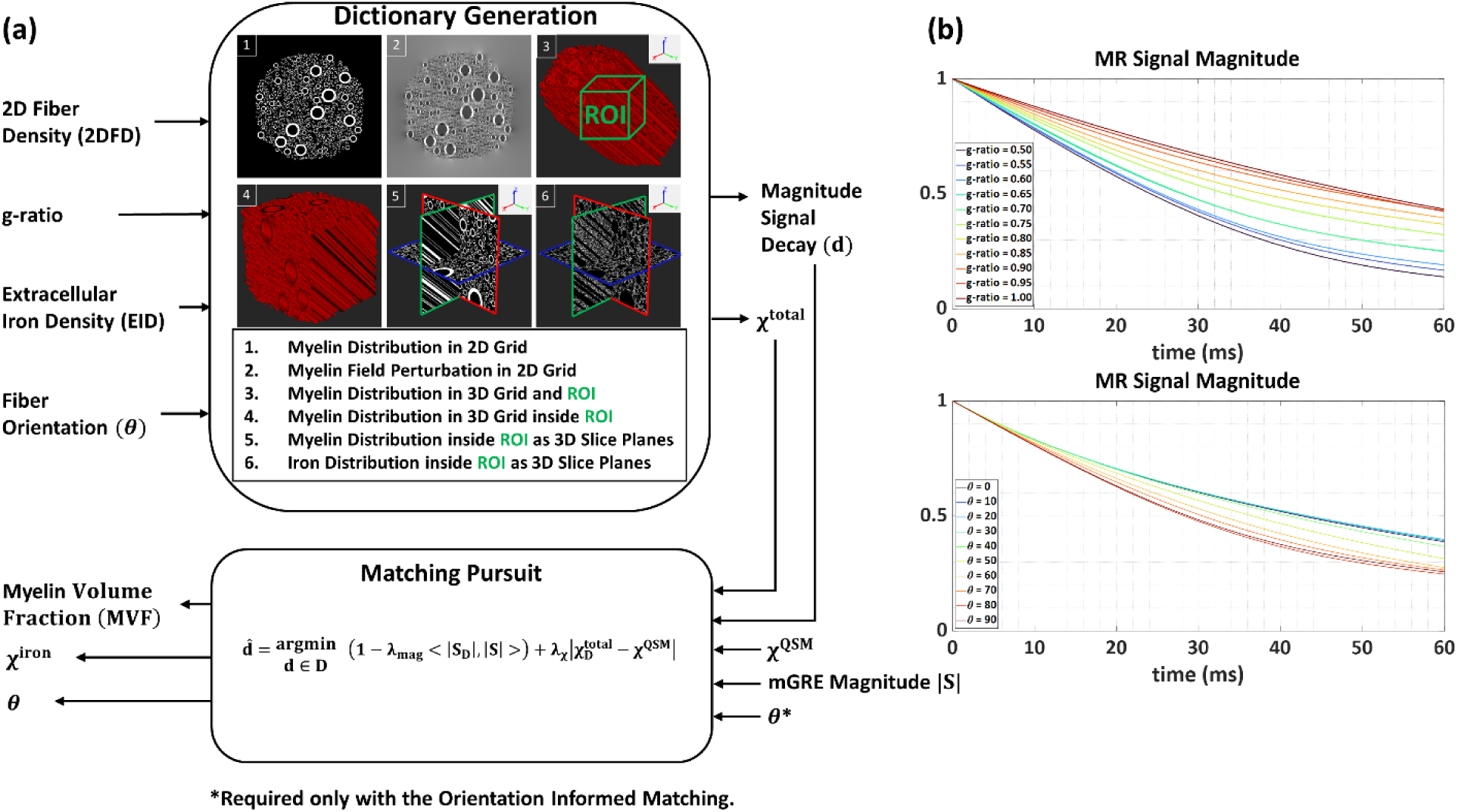
a) A detailed description of the dictionary generation and matching pursuit process. b) Example dictionary elements showing how the magnitude signal evolutions depend on the g-ratio and *θ* values.

It is important to note here that even though FVF and g-ratio are required to be decoupled for the simulation of the dictionary, however, the fitted values for them are not expected to be correct considering the mGRE signal is sensitive to total myelin content rather than fiber density and myelin thickness separately. Therefore, it is more convenient to claim a measure of the total myelin volume within each voxel. Consequently, the MVF value for each dictionary element can be computed as (Campbell et al., 2018):

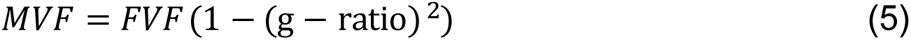

Furthermore, a bulk susceptibility value is also estimated for each dictionary element based on the volumetric myelin and iron susceptibility values and volume fractions. The voxel susceptibility originating from myelin content is calculated using the isotropic component as *χ*^*myelin*^ = *χ*_*iso*_ × *MVF* assuming that single orientation QSM measures the mean susceptibility of the susceptibility tensor; mean susceptibility is defined as the average of the eigenvalues of the susceptibility tensor (or 1/3 of the trace) (C. Wisnieff et al., 2013). Then, voxel myelin and iron susceptibilities are defined as *χ*^*myelin*^ = *χ*_*myelin*_ × *MVF* and *χ*^*iron*^ = *χ*_*iron*_ × *IVF*. Here, superscripts are utilized to refer to voxel quantities, whereas subscripts are reserved for volumetric quantities. Finally, the total susceptibility of each dictionary element is computed as:

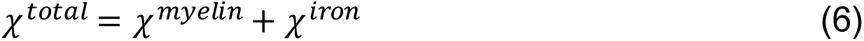

To create the overall dictionary that will be employed for MIMM, two approaches are adopted. First, as done in the conventional MR fingerprinting(Ma et al., 2013), the input parameters of the dictionary generation process are selected so that they will cover the parameter space uniformly. For instance, g-ratio is swept between 0.5 to 1 with a 0.05 step size. Secondly, the parameter space coverage is driven by a random sampling approach using a uniform distribution function again using the same range of parameters (Chen, 2011). Only the 2DFD is kept uniformly sampled to ensure comprehensive coverage of the parameter space while keeping the overall computation time short. After choosing the input parameters, the remaining steps are the same for both approaches. In this paper, the dictionaries generated with the explained approaches are referred to as the deterministic pursuit dictionary and the stochastic pursuit dictionary, respectively.

### 2.2. Stochastic Matching Pursuit of the Solution

To match the dictionary and the acquisition echo times, the magnitude signal evolution of each element is resampled to the acquisition echo times using a 5^th^-order polynomial interpolation of the natural logarithm of the dictionary signals. Then, the interpolated magnitude signals are normalized to have a unit Euclidian norm.

#### 2.2.1. Basic MIMM

The microstructural properties of each voxel are determined by an exhaustive search in the dictionary based on a novel similarity metric which is a linear combination of the traditional linear correlation between the dictionary and measured magnitude signal evolutions and the absolute difference between *χ*^*total*^of the dictionary elements and tissue susceptibilities (*χ*^*QSM*^) of the measurements. Linear correlation is widely used in MR fingerprinting for quantitative MR studies (Ma et al., 2013). *χ*^*QSM*^is the quantitative susceptibility value of each voxel estimated from the mGRE phase data. The idea to combine the magnitude signal evolution and QSM for quantitative imaging has previously been employed for MRI based oxygen extraction fraction (OEF) mapping (Cho, Spincemaille, Nguyen, Gupta, & Wang, 2021). The dictionary element that minimizes the following cost function is determined for each voxel *k*.

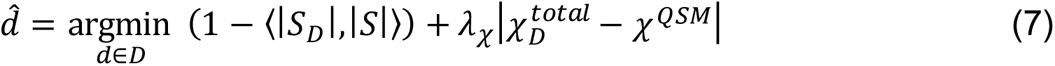

Here, |*S*_*D*_| and |*S*| are the normalized magnitude signal evolutions of the dictionary elements and the imaging voxel, respectively. *λ*_*χ*_is a weighting parameter that determines the relative weighting of the two terms. *D* is the dictionary and *d̂* is the dictionary element that minimizes the cost function. The corresponding *MVF*, *θ*, and *χ*^*iron*^values of *d̂* are then assigned to the voxel that is being processed to assemble the corresponding biophysical parameter maps.

#### 2.2.2. DTI Orientation Informed MIMM

Even though basic MIMM tries to tackle the fiber orientation dependence of the HCFM based MIMM by assuming *θ* is an unknown, it is known that in order to estimate the fiber orientation from mGRE measurements, data with at least 3 different sampling orientations is required (Wharton & Bowtell, 2013). Therefore, the *θ* values provided by the basic MIMM are expected to have high error, thereby also inducing errors in the estimated MVF. MIMM is improved here by incorporating a fiber orientation map obtained from a separate Diffusion Tensor Imaging (DTI) scan as:

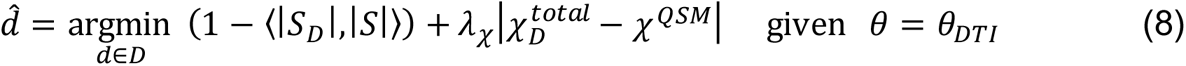

Here, the exhaustive search procedure to find the best matching element is limited to the dictionary elements whose orientation values (*θ*) match with the DTI fiber orientation (*θ*_*DTI*_). Before this procedure, the dictionary orientation values and the orientation values obtained from DTI are rounded to the nearest multiple of 5°. Moreover, orientation informed MIMM is applied only at the voxels where the fractional anisotropy (FA) value computed from the diffusion tensor (DT) is larger than 0.25 and the QSM value is lower than 0.1 ppm. This prevented using unreliable orientation values in the cortical gray matter where highly organized fiber tracts do not exist, and in basal ganglia, where DTI measurements can be erroneous due to the existence of high levels of iron.

#### 2.2.3. Atlas Orientation Informed MIMM

In the case that subject-specific WM fiber orientation is not available, we propose to use the orientation map obtained from a DTI brain atlas such as The International Consortium of Brain Mapping (ICBM) DTI-81 normative atlas (Mazziotta, Toga, Evans, Fox, & Lancaster, 1995; Mori et al., 2008). In this work, the atlas diffusion tensor was warped from the MNI ICBM-152 space to the subject mGRE space.

The MIMM algorithm is summarized in Figure 1a. Examples of dictionary magnitude signal evolution as a function of fiber orientation and g-ratio are shown in Figure 1(c-d).

## 3. Methods

Different aspects of the MIMM algorithm are evaluated utilizing Monte Carlo simulations, and experimental data from healthy subjects. The experimental myelin volume fraction maps are compared with myelin water fraction maps obtained with conventional T2-relaxometry (Nguyen et al., 2016; Nguyen et al., 2017). Furthermore, 3PCF is also implemented as described in (Nam et al., 2015) and the obtained MWF maps are compared with the MVF maps both quantitatively and qualitatively.

### 3.1. Simulation Grid Resolution and Size Optimization

As described in Section 2.1, in order to obtain the magnitude signal evolution of a dictionary element, a finite-size simulation volume needs to be created and the corresponding field perturbation distribution needs to be calculated. Considering the fact that an imaging voxel generally contains thousands of fibers, simulating a realistic voxel requires extremely high computation power and time. On the other hand, if we employ a relatively larger grid size of simulation than a single fiber, the resulting magnitude signal evolution may approximate the signal of a voxel that is substantially larger. In order to assess this idea, simulation volumes with varying resolutions and sizes were created (assuming the largest fiber radius is 10 *μm* (Graf von Keyserlingk & Schramm, 1984)) and the corresponding magnitude signal evolutions were computed. In Figures 2a and 2d, the created simulation volumes with varying grid resolution and sizes are shown, respectively. Figures 2b and 2e demonstrate the corresponding magnitude signal evolutions, and Figures 2c and 2f depict the corresponding computation time for each case. Figures 2b and 2c suggest that there is minimal change in the magnitude signal evolution between grid resolutions 256 and 512 even though the required computation time to obtain these signal curves is almost 5 times larger for the latter. Similar observations were made for grid sizes 50 *μm* and 100 *μm*. Therefore, for the rest of this study, all the dictionary elements are computed utilizing a 256^3^ grid at 50 *μm* resolution.

**Figure 2.**
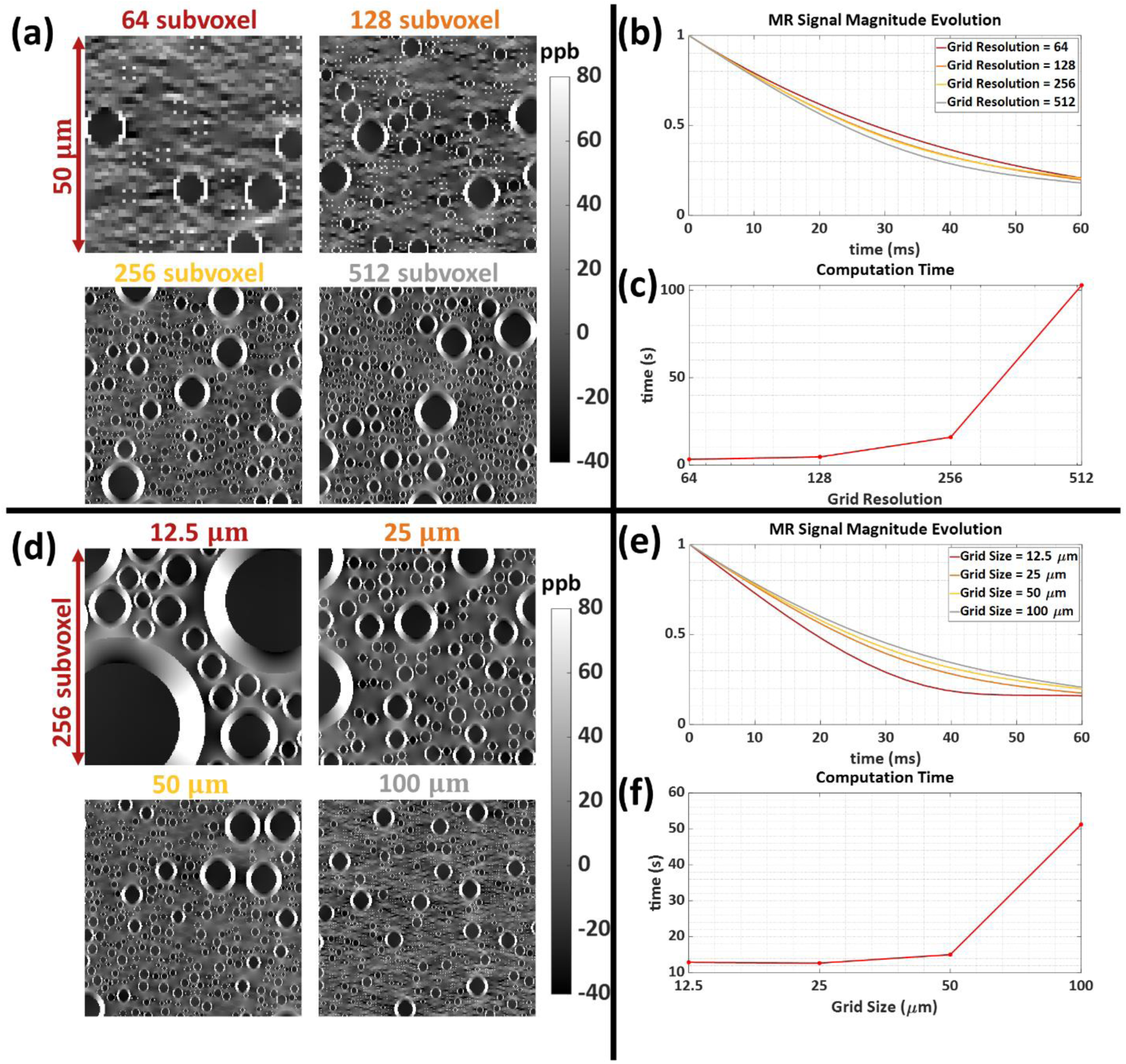
Comparison of different grid resolutions and grid sizes for the MR magnitude signal evolution simulations. a) Field perturbation distributions in 4 different resolution levels where the grid size is the same (50*μ*). b) Signal magnitude evolutions corresponding to 4 different resolutions. c) The time spent computing each signal magnitude evolution for each resolution. d) Field perturbation distributions in 4 different grid sizes where the grid resolution is the same (256 sub-voxel). e) Signal magnitude evolutions corresponding to 4 different grid sizes. f) The time spent computing each signal magnitude evolution for each grid size.

### 3.2. Monte Carlo Simulations

In order to evaluate the sensitivity of the MIMM approach to MRI signal noise, a separate set of data (*N* = 10000) was computed with random simulation parameters while keeping the constant parameters in Table 1 the same. For a given Signal-to-noise ratio (SNR) level, all the dictionary elements were corrupted with noise. Independent identically distributed (i.i.d.) Gaussian noise with standard deviation (s.t.d.) 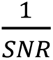 were added to both the real and imaginary parts of the signal evolutions before normalization (Note that the first echo magnitude is 1 before normalization). Moreover, Gaussian noise with s.t.d. 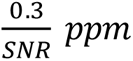 was also added to the *χ*^*total*^ values. Then, the noise corrupted signals and *χ*^*total*^ values were matched with the original dictionary as if it was a real measurement using various echo times and *λ*_*χ*_ values. The mean absolute error of the estimated MVF values *MVF*_*j*,*E*_ was computed as:

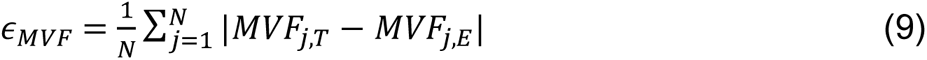

were *MVF*_*j*,*T*_were the true MVF value of the *j*^*t*ℎ^simulated data point. This was computed for the basic and the orientation informed MIMM strategies under different levels of noise. Moreover, it is also used to investigate the effect of the weighting factor *λ*_*χ*_ in Equations 7 and 8 and of the dictionary size.

### 3.3. MR Data Acquisition

Healthy subject data were acquired on a 3T MRI scanner (Prisma^fit^, Siemens Healthineers, Erlangen, Germany) using a 64-channel array head coil in ten volunteers (7 males, 3 females, age range: 25–51) following an IRB-approved protocol. Informed consent was obtained from each subject. The protocol for each scan was as follows:

(1) Whole-brain T1-weighted (T1w) scan using Magnetization Prepared Rapid Acquisition Gradient Echo (MPRAGE) with acquisition parameters: axial field of view FOV = 25.6 cm, phase FOV factor 100%, repetition time TR = 2300 ms, echo time TE = 2.26 ms, voxel size = 1 mm^3^ isotropic, readout bandwidth RBW = 200 Hz/pixel, flip angle FA = 8°, GRAPPA acceleration factor = 2, acquisition time = 5:21 min (176 slices).
(2) Monopolar 3D mGRE with acquisition parameters: FOV = 25.6 cm, phase FOV factor = 81.3%, TR = 41 ms, TE_1_/ΔTE = 2.2/3.25 ms, number of TEs = 7, voxel size = 1×1×2 mm^3^ (interpolated to 1 mm^3^ isotropic), RBW = 260 Hz/pixel, FA = 15°, slice partial Fourier factor = 0.875, GRAPPA acceleration factor = 2, acquisition time = 6:05 min (160 slices).
(3) FAST-T2 with acquisition parameters: 3-dimensional spiral acquisition with TR/TE = 7.6/0.5 ms, FOV = 25.6 cm, phase FOV factor=100%, sequence TR = 2000 ms; T2prep TEs = 0 (T2prep pulse turned off), 7.5, 17.5, 67.5, 147.5, and 307.5 ms, voxel size = 1.3×1.3×2 mm^3^ (interpolated to 1×1×2 mm^3^), RBW = 1042 Hz/pixel, FA = 10°, number of spiral leaves per stack = 32, acquisition time = 8 min (80 slices).
(4) 2D Spin Echo Echo Planar Imaging (SE-EPI) Diffusion Weighted Imaging (DWI) with acquisition parameters: number of diffusion encoding directions = 30, b-value = 1000 s/mm^2^, FOV = 24 cm, phase FOV factor = 100%, TR = 10000 ms, TE= 70 ms, voxel size = 1.9×1.9×2.5 mm^3^, RBW =1562 Hz/pixel, slice partial Fourier factor = 0.75, GRAPPA acceleration factor = 2, acquisition time = 5:40 min (60 slices).

### 3.4. Data Processing

#### 3.4.1. ROI Segmentations

For regional MWF and MVF analysis, a total of 15 ROIs were selected for each subject: 5 subcortical GM ROIs (Globus Pallidus, Putamen, Caudate, Thalamus, Hippocampus) were extracted from *T*_1_*w* images using FreeSurfer(Fischl, 2012); and 10 WM ROIs (Genu, Splenium, and Body of Corpus Callosum, External Capsule, Anterior Limb of Internal Capsule, Optic Radiation, Superior Longitudinal Fasciculus, Superior Corona Radiata, Cingulate Gyrus, and Cerebro Spinal Tract) were chosen from the WM parcellation map provided by the DTI-81 atlas (Mori et al., 2008).

#### 3.4.2. DTI Image Reconstruction

First, the susceptibility induced *B*_0_ off-resonance map is estimated from mGRE phase data using a nonlinear fitting algorithm (T. Liu et al., 2013) and spatially unwrapped (Cusack & Papadakis, 2002). Then, DWI images were corrected for geometric distortions using the estimated off-resonance map and FSL FEAT algorithm (Jenkinson, Beckmann, Behrens, Woolrich, & Smith, 2012; Woolrich, Ripley, Brady, & Smith, 2001). Finally, Diffusion Tensor images were reconstructed using the FSL DTIFIT algorithm (Behrens et al., 2003).

#### 3.4.3. QSM Image Reconstruction

QSM images were reconstructed from the same field used for DTI image correction. The background field is removed using the projection onto dipole fields (PDF) algorithm (T. Liu et al., 2011) and QSM is reconstructed using Morphology-Enabled Dipole Inversion (MEDI) with global cerebrospinal fluid (CSF) referencing (A.V. Dimov et al., 2022; J. Liu et al., 2012; Z. Liu, Spincemaille, Yao, Zhang, & Wang, 2018).

#### 3.4.4. Myelin Content Image Reconstructions

The macroscopic field inhomogeneity effects in the mGRE magnitude data are corrected using the library-driven voxel spread function approach (Y. Liu et al., 2021; Yablonskiy, Sukstanskii, Luo, & Wang, 2013). The MIMM algorithm was implemented in MATLAB R2020a (Mathworks, Natick, Massachusetts, USA), and all the in vivo data were processed using the same implementation. Furthermore, the 3PCF algorithm proposed by Nam et al. (Nam et al., 2015) was also implemented to estimate the MWF map using the mGRE data. The implementation details provided in (Nam et al., 2015) were followed in addition to some additional smoothing introduced before fitting to suppress noise, due to the lower number of echoes available. Finally, MWF from FAST-T2 was extracted as described in (Nguyen et al., 2016).

For the in vivo data, *λ*_*χ*_ was determined using L-curve analysis (Hansen, 1992) by balancing the two error terms in Equations 7 and 8 in a representative subject. It was also found this balancing can be limited to a subset of the data in a subject: In this work, *λ*_*χ*_was determined by averaging the two error terms obtained within all the voxels in the 10 central slices.

Selecting the dictionary size poses a tradeoff between the goodness of the match (it is possible to find a better match in the dictionary with a larger size) and the time required for the matching pursuit process. In a representative subject, the MVF map was obtained with different dictionary sizes and the average minimum matching error together with the image entropy (Pal & Pal, 1991) were computed over the whole brain white matter. Image entropy level was employed to quantify the detail in the image features.

#### 3.4.5. Image Registrations

Several registrations between different subject image spaces and to/from ICBM −152 space were required during data processing. All the registrations can be summarized here:

(1) A linear transformation (affine transform) from subject DTI space to mGRE space was estimated using FSL Linear Image Registration Tool (FLIRT) (Jenkinson, Bannister, Brady, & Smith, 2002), the b-value = 0 image from DTI dataset and combined T2*-weighted (T2*w) magnitude image of the mGRE and the FA map was brought to mGRE space. The first principal eigenvector of the reconstructed DTI images (*V*_1_) was registered to the mGRE space using the estimated linear transformation and FSL *vecreg* command to estimate *θ* maps.
(2) Nonlinear transformation (affine transform and warp field) was estimated from ICBM-152 space to individual subject T1 space using ANTs *SyN* algorithm (Avants, Tustison, & Song, 2009). Then, a linear transformation from subject T1 space to mGRE space was estimated again using ANTs. The ICBM-81 DTI atlas was brought to the mGRE space using the computed transformations as a single-step transformation. The first principal eigenvector (*V*_1_) of the registered ICBM-81 DTI atlas and then the corresponding *θ* maps were calculated.
(3) The linear transformation from subject FAST-T2 space to mGRE space was estimated using FSL FLIRT to bring the estimated reference MWF maps to mGRE space.
(4) Finally, ROI segmentation masks that reside on subject T1 and ICBM-81 DTI atlas spaces were brought to subject mGRE space with FSL FLIRT and ANTs using the computed linear and nonlinear transformations, respectively.

A desktop PC (CPU: Intel i7-5820k, 3.3 GHz; 64 GB RAM) was used for all the data processing steps.

## 4. Results

Figure 3a shows example MVF maps obtained using the basic matching pursuit with both the deterministic and stochastic versions, as well as using two dictionary sizes. MVF maps obtained with stochastic matching are visibly smoother and present less “salt-and-pepper-noise” compared to those obtained using deterministic matching. No obvious improvement with the increased dictionary size was observed, even though the larger dictionary increases the computational cost of matching by a factor of 4. To visualize the source of this significant difference in the results, we projected the input parameters of 2 example dictionaries (deterministic pursuit dictionary with size 12540 and stochastic pursuit dictionary with size 12000) onto the (g-ratio, *θ*) plane (Figure 3b). Even though the total number of dictionary elements is similar, the parameter coverage of the stochastic pursuit dictionary is improved in certain subspaces. Furthermore, in Figure 3c all the dictionary elements are shown g-ratio − *θ* − IVF parameter space to visualize the 3D space coverage. Even though it is difficult to see the difference between the two dictionaries as in the 2D case, it can be seen that the total number of elements is similar.

**Figure 3.**
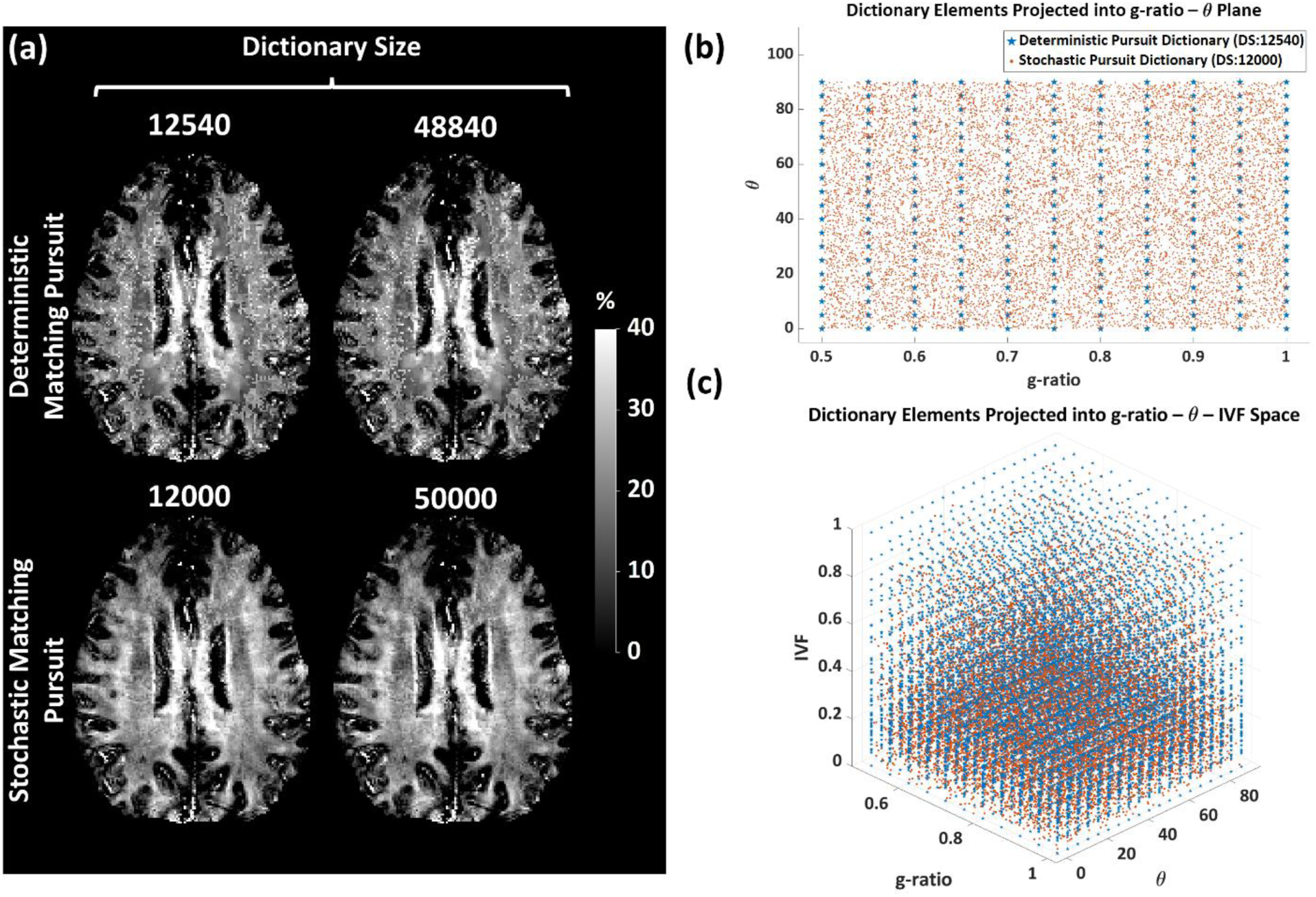
Comparison of the deterministic and stochastic matching pursuit (MIMM) results. a) Example MVF maps obtained with deterministic matching pursuit and MIMM with different dictionary sizes. Basic MIMM is utilized in this example. b) Projection of all the dictionary elements utilized for the two pursuits onto the g-ratio − *θ* plane. c) Projection of all the dictionary elements onto g-ratio − *θ* − IVF parameter space.

Figure 4a shows two L-curve results for different subjects. *λ*_*χ*_ = 0.015 was approximately located at the corner of the L-curve for both subjects and then used for all the further results. Example MVF maps using both the basic and orientation informed MIMM for 3 different values of *λ*_*χ*_ are demonstrated in Figure 4b. Additionally, Figure S1 presents the results of the Monte Carlo experiments to evaluate the performance of the MIMM based quantification of the microstructural properties with different *λ*_*χ*_ values. It demonstrates the mean absolute errors of the 3 output parameters of the MIMM algorithm for both basic and orientation informed MIMM strategies with both deterministic pursuit and stochastic pursuit dictionaries. The plots show that mean absolute errors increase with decreased SNR levels as expected. Furthermore, for most of the results, the accuracy is higher when orientation informed MIMM strategy is utilized. The error levels are relatively constant for a large range of *λ*_*χ*_ values except between 10^−4^ and 10^2^, where an optimal value of *λ*_*χ*_ maximizes the accuracy of the fitting. However, this value is SNR-dependent.

**Figure 4.**
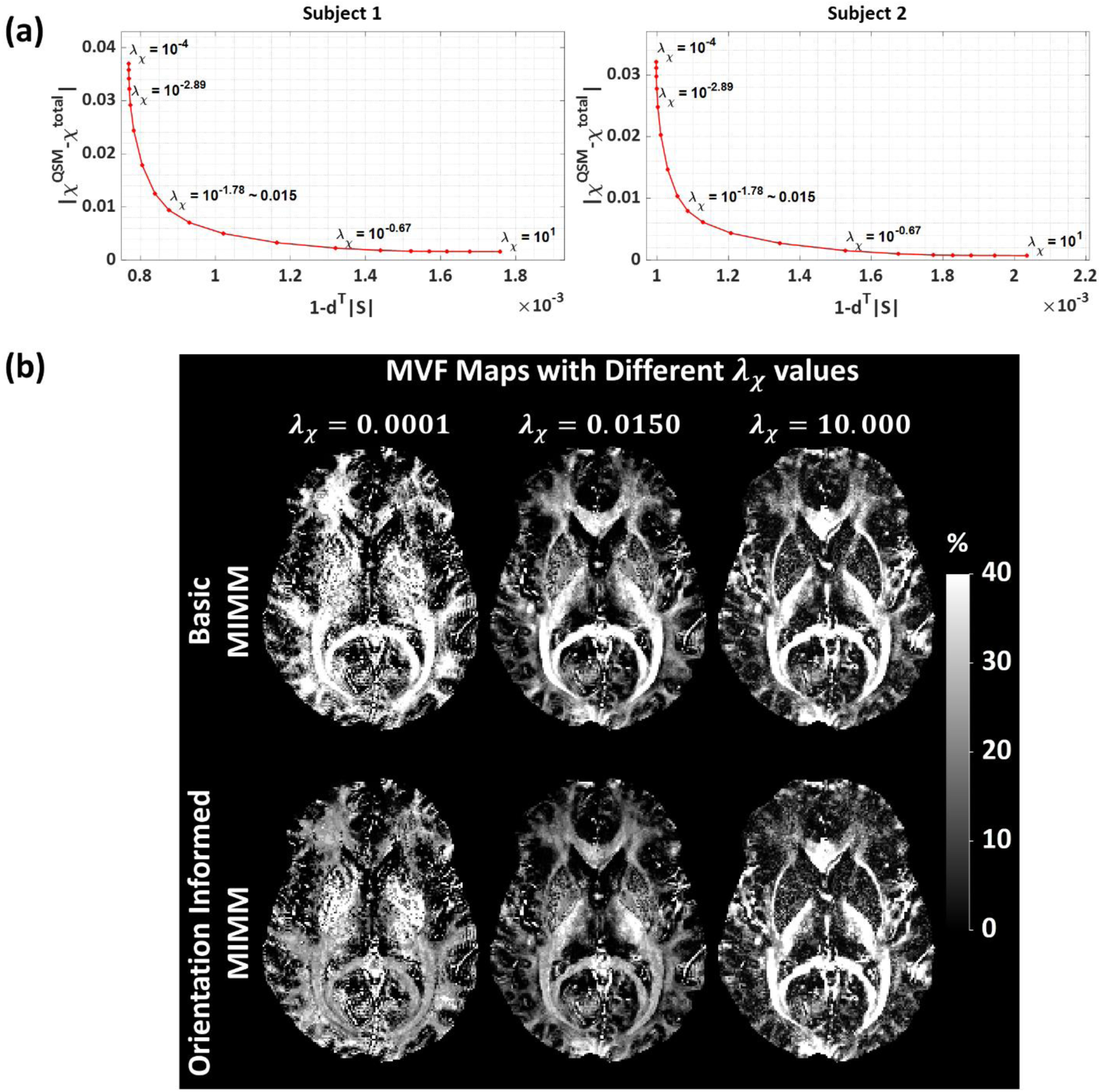
Analysis regarding the choice of the optimal weighting parameter *λ*_*χ*_. a) L-curve analysis results for 2 different subjects using basic MIMM. The values demonstrated in the plots are obtained by averaging the errors obtained with the given *λ*_*χ*_among all the voxels in 10 central slices. b) MVF maps obtained using basic and orientation informed MIMM using DTI and the dictionary of size 20000 with different *λ*_*χ*_ values.

Figure 5a shows a comparison between various dictionary sizes. From dictionary size 20000 to 80000, the minimum matching error decreases by 6%, and the image entropy increases by only 1%. These results suggest that the increase in the matching performance and the change in the image features are subtle after a dictionary size of 20000 even though the dictionary size and consequently the matching time increases by a factor of 400%. Hence, a dictionary size of 20000 was employed for all the results hereafter unless otherwise stated. Example MVF results using both the basic and orientation informed MIMM for 3 different dictionary sizes are shown in Figure 5b. Figure S2 shows the Monte Carlo simulation results regarding the dependence of the accuracy of the MIMM algorithm on the dictionary size. It is seen that after an approximate dictionary size of 10000, changes in the accuracy are below 4%.

**Figure 5.**
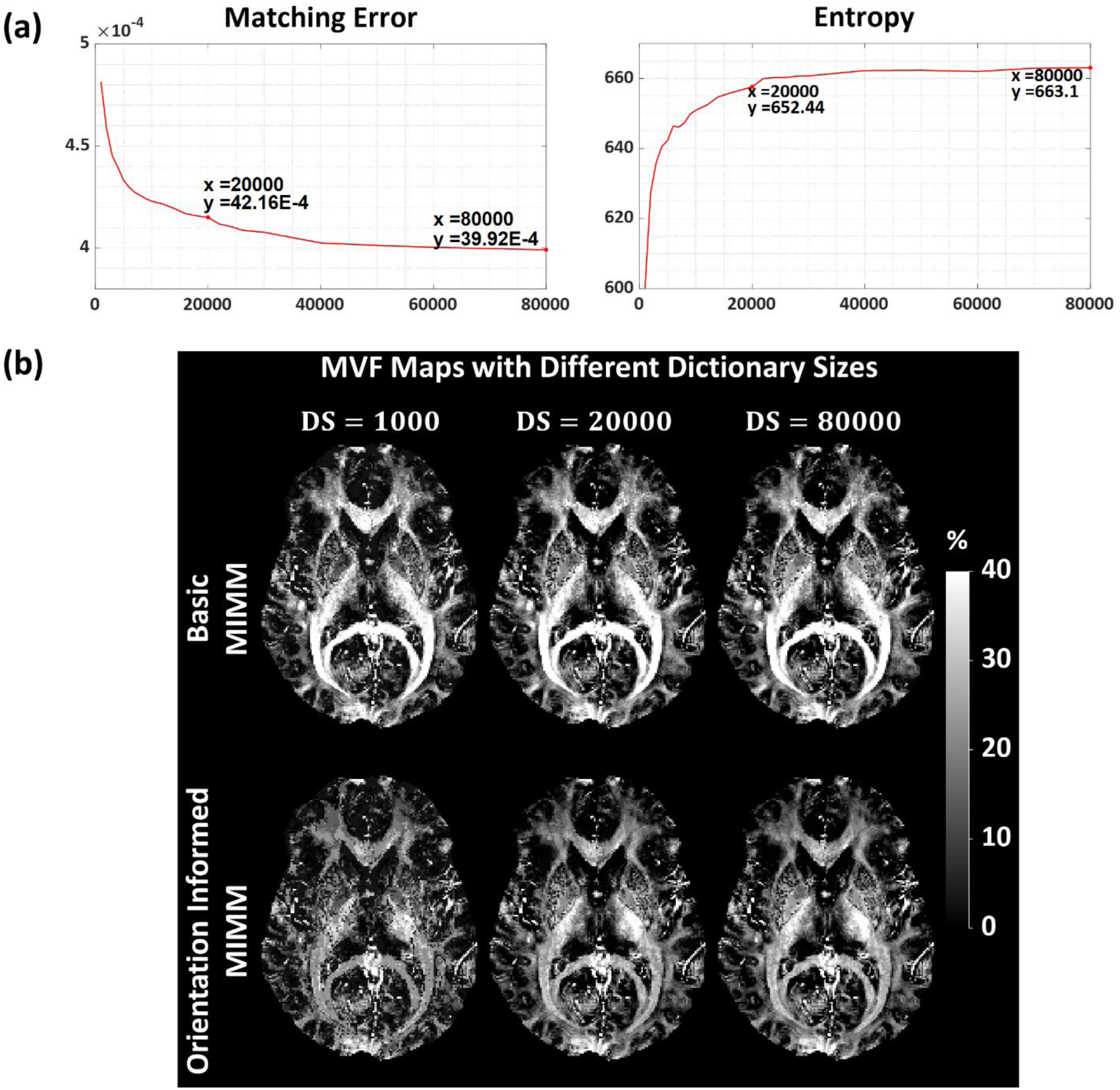
Analysis regarding the choice of the optimal dictionary size. a) Average minimum fitting error and image entropy values computed over the white matter for an example subject for basic MIMM. b) MVF maps obtained using basic and DTI orientation informed MIMM using different dictionary sizes using *λ*_*χ*_ = 0.015.

Four axial slices of the obtained MVF maps in a single subject using the basic MIMM strategy, DTI orientation informed strategy, and atlas orientation informed strategy are shown in Figure 6 and Figure S4 along with the corresponding T1w images, QSM maps, MWF maps obtained with 3PCF from the mGRE data, MWF maps obtained with T2-relaxometry from the FAST-T2 data and FA maps. All MVF maps correlated well visually with the reference FAST-T2 MWF maps. Compared to 3PCF MWF maps, MVF maps display a lower level of noise and image artifacts. The globus pallidus is visibly overestimated in the 3PCF MWF maps compared to the MVF maps (red arrows). Orientation informed MIMM reduces some of the visible overestimations in the main fiber tracts compared to basic MIMM (green arrows). However, overestimation is still present in certain tracts like the posterior limb of the internal capsule (yellow arrows) that correspond to the fiber tracts oriented in the superior-inferior direction (blue/purple regions in the FA map).

**Figure 6.**
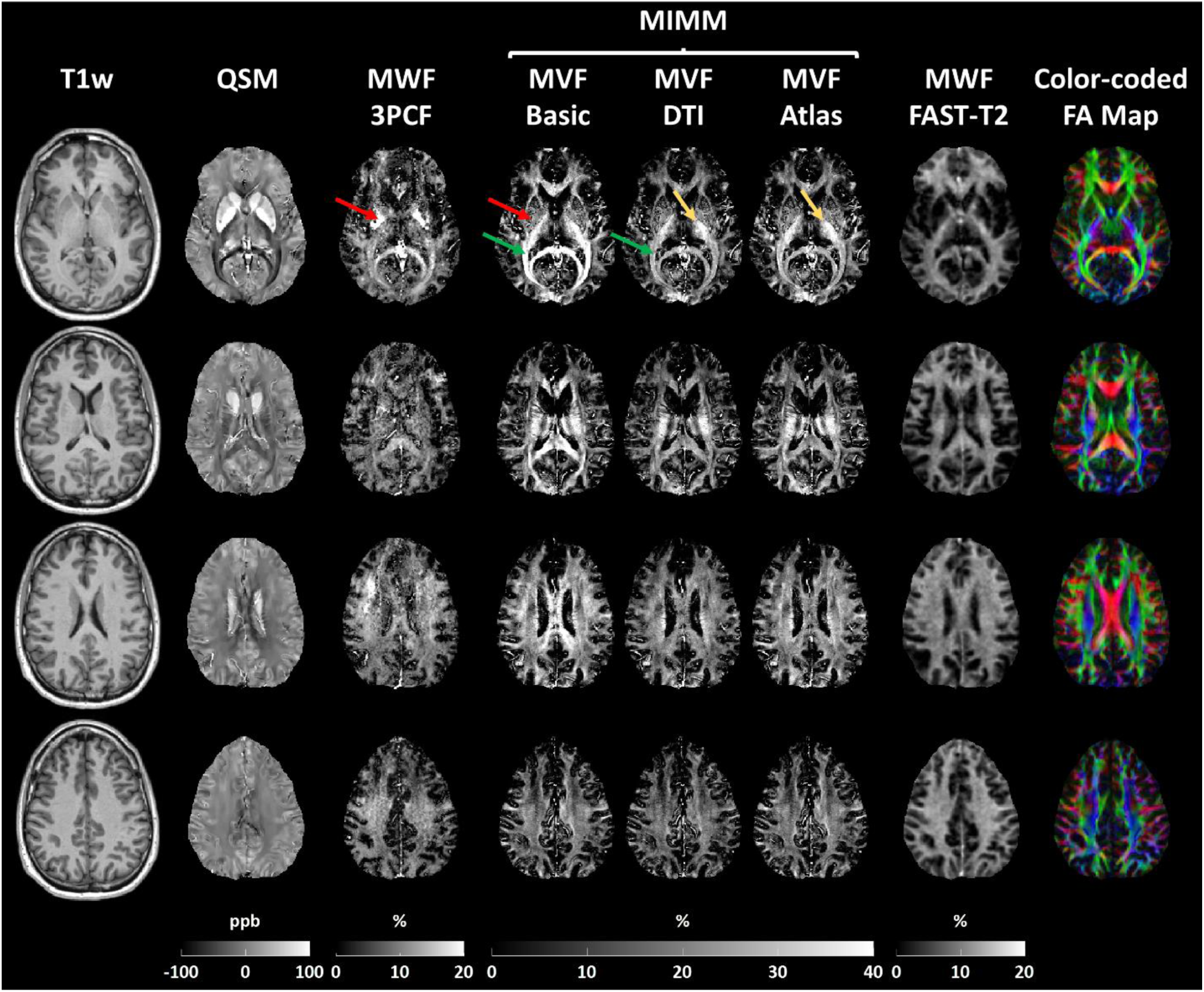
Example results from a single subject. From left to right: T1-weighted images (T1w), QSM maps, MWF maps obtained with 3 pool complex fitting (3PCF), MVF maps obtained with basic MIMM, DTI orientation informed MIMM, and atlas orientation informed MIMM, MWF maps obtained using T2-relaxometry from the FAST-T2 data, and FA maps obtained from diffusion tensor images. All images are registered to the QSM space.

Figure 7 shows the results from the ROI based linear correlation analysis between the MVF maps obtained with all 3 MIMM strategies and the FAST-T2 MWF maps, as well as that between 3PCF MWF and FAST-T2 MWF for comparison. All 3 MVF maps show a significant correlation with FAST-T2 MWF with a correlation of r ≈ 0.73-0.75 whereas the 3PCF MWF map shows a significant correlation with r = 0.34. Subject-specific linear correlation analysis results for each method are presented in Figures S4-S8.

**Figure 7.**
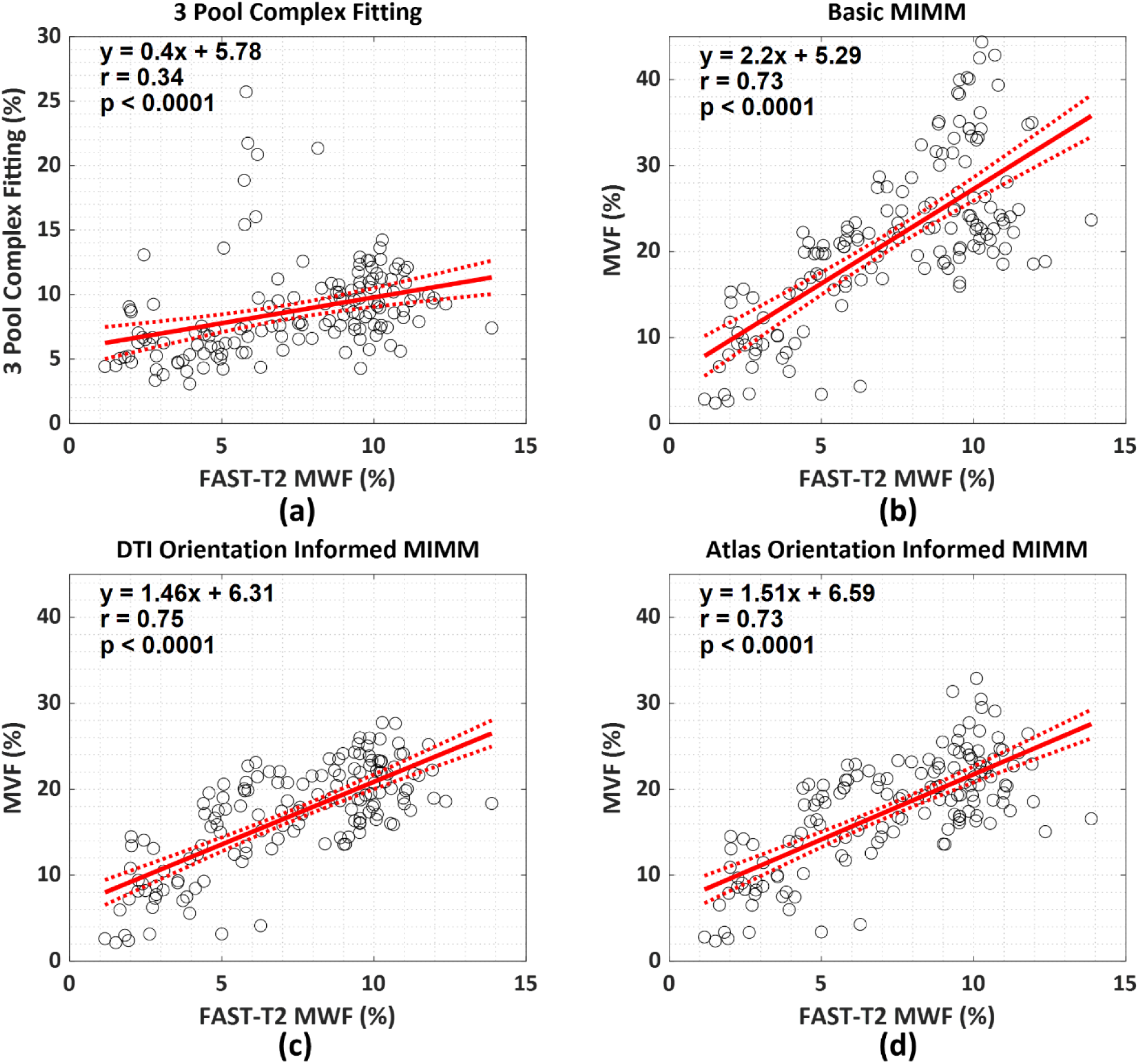
Linear correlation analysis results between the FAST-T2 based MWF maps and a) the MWF maps obtained using the 3 pool complex fitting, the MVF maps are acquired from maps obtained using b) basic MIMM, c) DTI orientation informed MIM, and d) atlas orientation informed MIMM.

A comparison of the orientation maps in a single subject between the ICBM DTI-81 atlas based approach and the subject DTI orientation based map is presented in Figure 8. The two orientation estimates as well as the corresponding MVF maps are compared using linear regression and Bland-Altman analysis of the ROI averages in Figure 8b.

**Figure 8.**
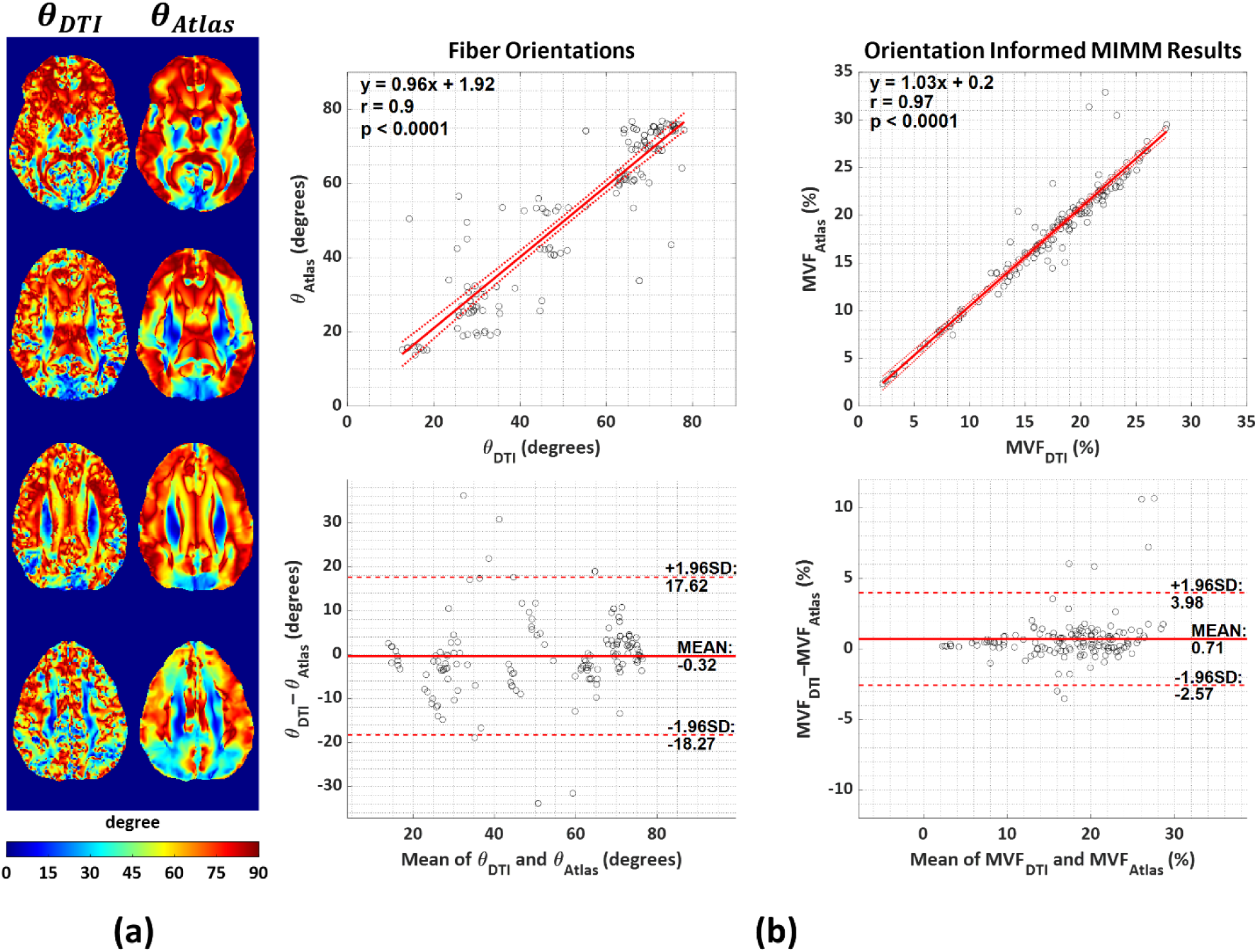
a) Comparison of the orientation maps extracted from DT images (*θ*_*DTI*_) and ICBM DTI-81 atlas (*θ*_*Atlas*_). b) Results of the linear correlation and Bland-Altman analysis between the ROI orientation angle values (left column) and the ROI MVF values obtained using orientation informed MIMM using the corresponding orientation maps.

Figure 9a presents example iron susceptibility distribution maps obtained with 3 MIMM strategies for an example subject. Average *χ*^*iron*^values across all the subjects within different cortical gray matter parcellations are shown in Figure 9b. The voxel-wise distribution of the MVF and *χ*^*iron*^for all 10 subjects are presented in Figure 10 as violin plots for three major brain regions (cerebral white matter, cortical gray matter, and deep gray matter). In order to suppress voxel-wise noise effects in this plot, the parameter maps were blurred with a Gaussian kernel of size 5×5×5 beforehand.

**Figure 9.**
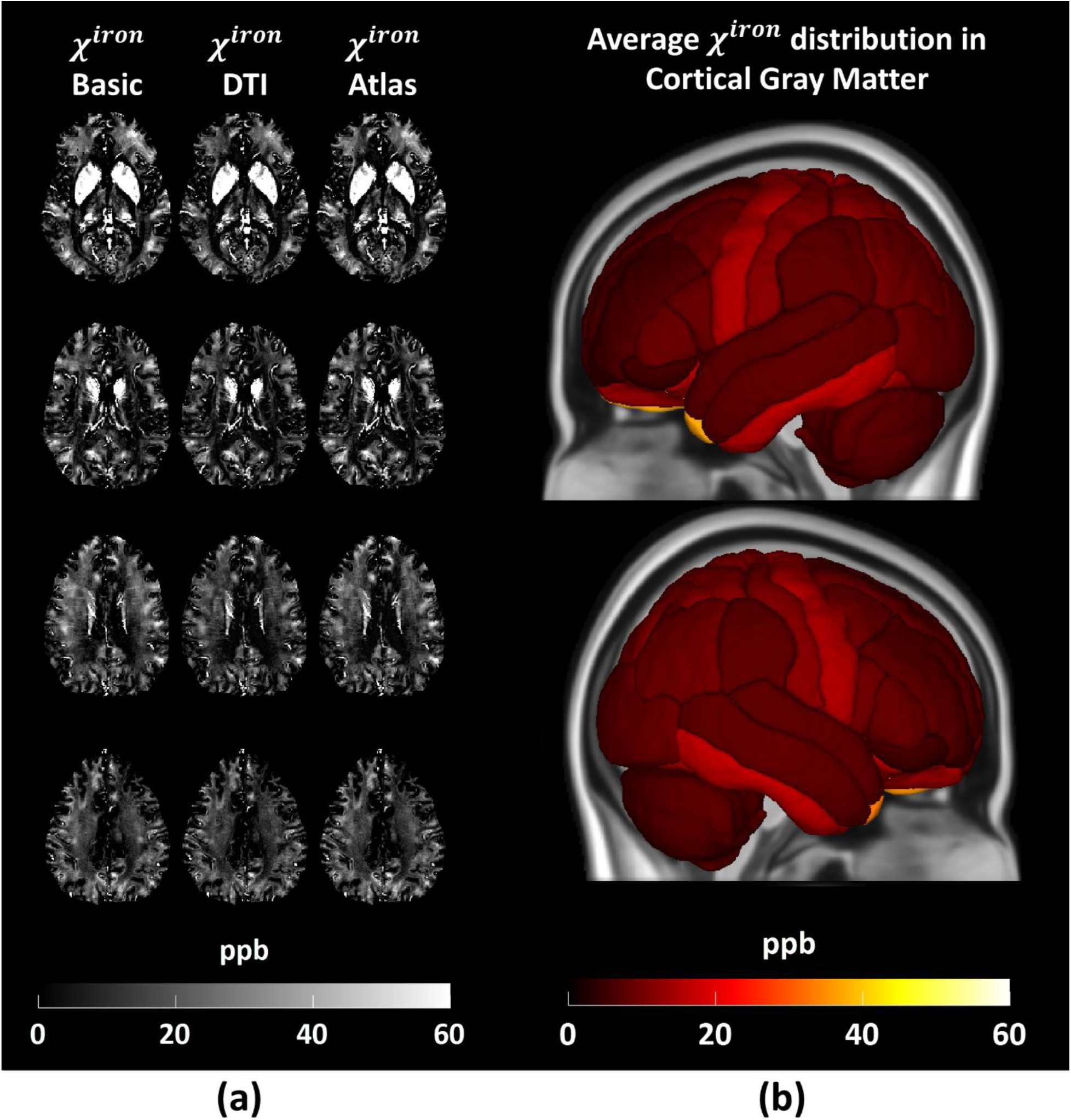
Example results of the *χ*^*iron*^maps. a) *χ*^*iron*^ maps obtained from a single subject. From left to right: *χ*^*iron*^maps obtained with basic MIMM, DTI orientation informed MIMM, and atlas orientation informed MIMM. b) Average *χ*^*iron*^values across all subjects within different cortical gray matter parcellations.

**Figure 10.**
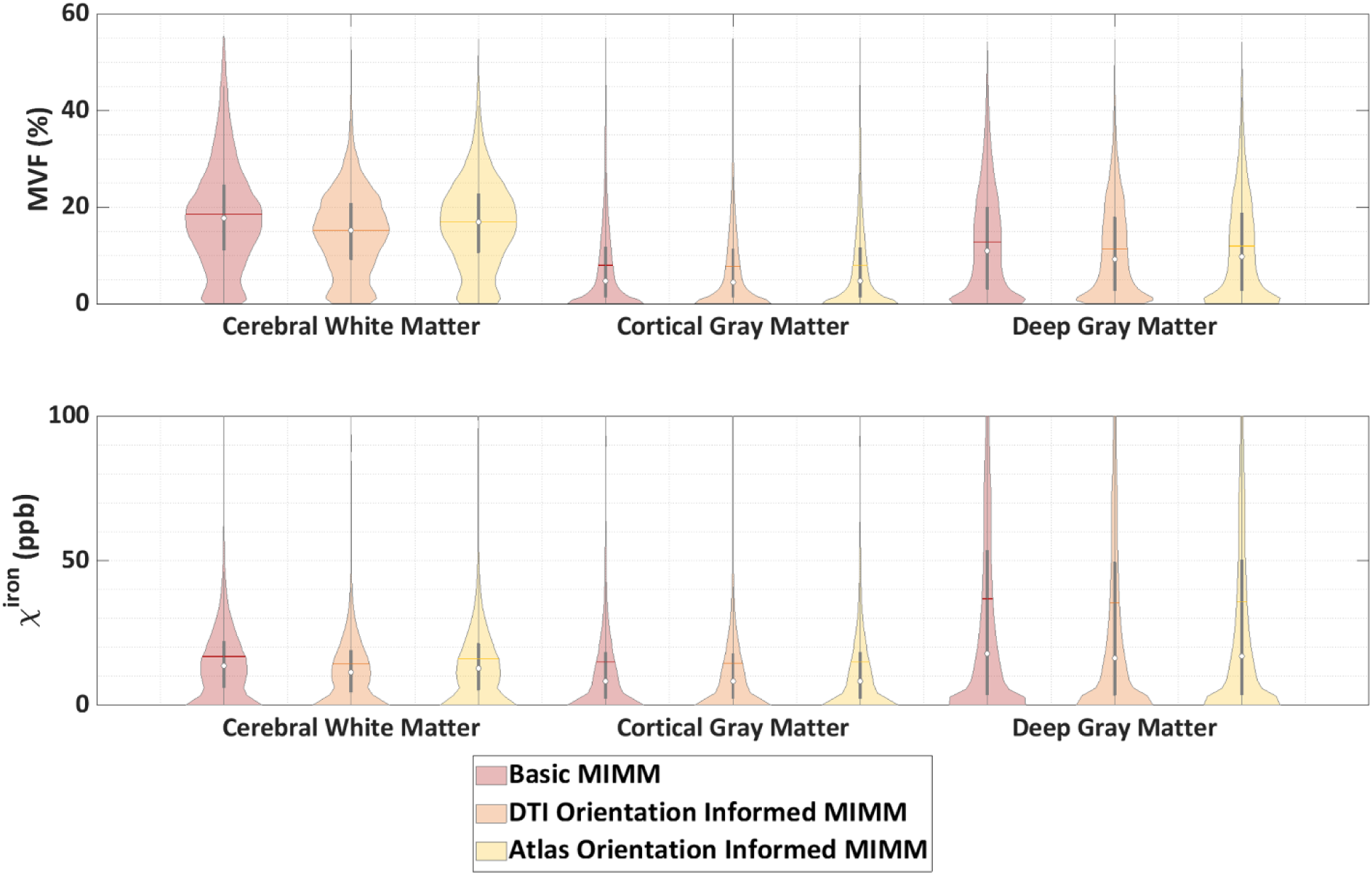
Violin plots showing the distribution of the voxel values in MVF (top row) and *χ*^*iron*^ (bottom row) maps combined over all 10 subjects for 3 major brain regions.

In order to evaluate the reproducibility capability of the results with MIMM, the MVF maps of the same subject are computed 8 times independently with 8 different dictionaries with dictionary size 10000 and *λ*_*χ*_ = 0.015 utilizing orientation informed MIMM. The reconstructed maps and the voxel-wise mean and standard deviation images are depicted in Figure S3.

## 5. Discussion

In this study, we have demonstrated the feasibility of using an HCFM based modeling of the myelin sheaths and the inclusion of iron sources for the quantification of myelin in brain tissue. Better correlation with the reference MWF maps and better visual quality were obtained using the MIMM algorithm compared to the prior 3PCF. Fiber orientation helps myelin mapping accuracy at white matter tracts, and atlas information seems sufficient, providing similar results as subject-specific DTI orientation maps. Finally, paramagnetic iron-specific susceptibility distributions are demonstrated as an additional output of the MIMM algorithm.

MIMM improves myelin mapping by better modeling the mGRE signal of white matter in the following aspects compared to prior methods, which is consistent with findings from improving QSM biophysics modeling (A. V. Dimov et al., 2015; A. V. Dimov et al., 2018; Kee et al., 2017; Wen et al., 2021). First, realistic HCFM is used to model the field perturbation distribution due to the myelin content of tissues and the corresponding magnitude decay curves. Second, the brain iron is modeled as an additional susceptibility source that was not explicitly accounted for in the previous methods. Finally, local QSM values are utilized for voxel-wise myelin and iron quantification rather than the non-local phase.

Stochastic matching pursuit provides effective filtering of signals not modeled in the dictionary. This study considers 2 effective water pools (intra-extracellular and myelin): myelin water as a bound water pool that is trapped between the lipid bilayers of the myelin sheath demonstrates much faster decay and the water pools inside and outside the cells. The CSF water pool that is included in a lot of the previous MWF works (Mackay et al., 1994; A. L. MacKay & Laule, 2016; Nguyen et al., 2016) demonstrates negligible decay in most of the mGRE acquisitions with relatively short echo times (T2_CSF_ > 600 ms) (Meyers, Kolind, & MacKay, 2017). Therefore, the CSF water pool can be effectively ignored in constructing the dictionary.

The use of random grid sampling of the dictionaries in stochastic matching pursuit is very useful for accurate myelin mapping at a given SNR level, as shown in simulations (Figure S1). In vivo, smoother MVF maps with lower apparent noise (Figure 3a) were obtained using the stochastic pursuit dictionary. Stochastic pursuit reduces the quantization mismatching errors due to the large gaps in the sampled parameter locations in the deterministic pursuit dictionary as seen in Figure 3(b-c). Particularly, the stochastic pursuit dictionary projected onto the g-ratio − *θ* plane provides smaller gaps or much denser parameter distribution although the dictionary sizes are similar. Figure 3b shows an almost continuous sampling of the g-ratio with the stochastic pursuit dictionary whereas a fixed quantization level of 0.05 in the deterministic pursuit grid dictionary is responsible for the apparent quantization errors in Figure 3a. Another benefit of the random sampling strategy for dictionary generation is the fact that the dictionary size can be increased in any desired fashion instead of the exponential growth of a conventional dictionary that arises by increasing sampling density in each parameter direction. This provides the flexibility that an optimum dictionary size can be determined more easily from a linearly increasing dictionary size search range as illustrated in Figure 5 and Figure S2. One drawback of the utilization of stochastic pursuit dictionaries is the intrinsic uncertainty introduced in the results by the different randomness of the dictionary. This can be observed in Figure S3 as a certain level of variation is observed in the results reconstructed with different dictionaries of the same size. Even though the visual differences are minimal between different results, there is a voxel-wise standard deviation that goes as high as 2% in the white matter. This can be an issue for reproducibility unless the same dictionary is used for the reconstruction of different cases, or a fixed random seed is utilized for the dictionary generation. This observation requires further investigation in the future.

The L-curves shown in Figure 4a demonstrate well the behaved selection of the weighting strength. Significant changes in the image contrast and artifacts were observed in Figure 4b as *λ*_*χ*_ increased. First of all, for low *λ*_*χ*_, the magnitude term in Equations 7 and 8 dominates during the matching pursuit process which diminishes the ability to separate myelin and iron. The basal ganglia, especially globus pallidus, are assigned very high MVF values. High *λ*_*χ*_values, on the contrary, lead to overestimated MVF values in the fiber tracts where the QSM values are very low (negative and high in magnitude) even with orientation informed MIMM and noisy texture in the remaining areas. MVF overestimation cannot be addressed in this case since orientation information is employed in the magnitude term. The intermediate values provide a balance between the two terms and reduce the errors using the complementing information arising from the two terms.

The MIMM algorithm addresses two major disadvantages in the prior 3PCF approach for MWF mapping: the lack of information regarding the separation of myelin and iron and the high number of model parameters. The former creates an artificial overestimation of the myelin content in regions with high iron deposition such as basal ganglia while the latter causes noisy parameter maps due to the intrinsic ill-posedness of the fitting process with a high number of fitting parameters. The inclusion of iron in the biophysical model and QSM in the matching provides a framework to separate the magnitude decay due to myelin and iron. Additionally, the 3PCF approach also requires a much longer computation time which can be a detrimental factor in clinical settings. For instance, the fitting of a single subject took ∼3 hours with our computation setting whereas the MIMM algorithm took approximately 4 mins. Moreover, utilization of the HCFM further accounts for the orientation effects in the white matter fiber tracts. The matching pursuit process with the biophysical modeling provides MVF maps with higher visual quality since the effective number of parameters that are solved is lower.

In Figure 7, the linear correlation with the reference FAST-T2 MWF obtained with all of the three MVF quantification methods was higher compared to that of the 3PCF MWF maps. This is in line with the improved qualitative performance shown in Figure 6. The surprising result here is that utilization of the orientation informed MIMM seems to provide no quantitative improvement over the basic MIMM. This is in contrast to the lower mean absolute errors observed in the simulation (Figure S1) and the better visual consistency with the FAST-T2 MWF map in Figure 6. Here, it should be noted that when orientation informed MIMM is utilized, not only does the vertical dispersion in the data points in Figure 7 decrease but the slope of the best-fitting line also decreases. These two parallel decreases both in the slope and data dispersion probably cancel each other out and produce a similar correlation coefficient. This can be explained considering most of the ROIs chosen in this study belong to the major white matter fiber tracts and the MVF values in these tracts decrease consistently when the orientation informed MIMM is utilized. A similar correlation does not mean similar accuracy since the correlation coefficient between two vectors will stay the same if one vector is scaled by a constant whereas the accuracy will change. Even though we showed that orientation informed MIMM provides better estimates of the MVF, basic MIMM has its practical advantage where it does not require any orientation map and still provides decent MVF distributions except for the overestimated values in the major fiber tracts. The promising result in Figure 8 is that almost identical MVF can be obtained using an atlas based fiber orientation distribution that is registered to the mGRE space rather than the orientation maps obtained from a separate DTI acquisition. The availability of a T1 weighted scan, which is often acquired in clinical protocols, can enhance image registration that is required for the atlas orientation informed MIMM. In a scenario where it is not available, direct registration of the atlas based diffusion tensor to the mGRE space can be realized but the accuracy might get lower. The MVF made available from standard mGRE using MIMM processing described in this work would allow routine study of myelin in various neurological disorders, including multiple sclerosis (Dobson & Giovannoni, 2019; Eskreis-Winkler et al., 2015; Cynthia Wisnieff et al., 2015) and amyotrophic lateral sclerosis(S. Kolind et al., 2013; Schweitzer et al., 2015).

A natural byproduct of the MIMM method is the *χ*^*iron*^distribution that demonstrates the concentration of iron in different brain regions. Even though this was not the main focus of this study, mapping the susceptibility distribution due to iron is also a clinically relevant task. Some examples of clinical interest in iron susceptibility mapping are the paramagnetic rim lesion detection in MS patients (Marcille et al., 2022) and motor cortex iron quantification in Amyotrophic Lateral Sclerosis (ALS) patients (Adachi et al., 2015). In Figure 9a, the *χ*^*iron*^ maps of the healthy subject are almost identical for the three matching strategies. This shows that orientation informed MIMM does not substantially impact the *χ*^*iron*^ distribution. This is likely due to the isotropic nature of the iron-related susceptibility sources. Figure 9b shows the non-uniform iron concentration in different gray matter regions of healthy subjects. Interestingly, in cortical gray matter, the motor cortex shows slightly higher iron concentrations compared to other regions, consistent with the literature (Kwan et al., 2012). After the separation of the myelin and iron content in the voxels, it is possible to evaluate their individual distributions in different regions of the brain. The violin plots in Figure 10 are provided as an example visualization of this kind of evaluation. MVF values have higher mean values in the white matter compared to cortical gray matter and deep gray matter as expected. Deep gray matter regions seem to have significant myelin content as well. In terms of iron, on the other hand, deep gray matter regions seem to have the highest content. The mean iron content in white matter and cortical gray matter regions seem to be close whereas the median iron content in white matter seems to be higher than cortical gray matter. The highest positive susceptibility source content in the cortical regions was observed to exist in the superficial white matter in the previous studies (S. Lee, Shin, Kim, & Lee, 2023) which can explain the current results. The results with different MIMM strategies, however, do not demonstrate an obvious difference in the distributions.

Though the use of an orientation prior in MIMM improves the myelin quantification in most of the white matter fiber tracts such as corpus callosum and optic radiation (green arrows in Figure 6), it fails to correct the overestimation in some fiber tracts such as the posterior limb of the internal capsule (yellow arrows in Figure 6). The main reason for the overestimation in those areas in the first place is the strong negative susceptibility effects in these fiber tracts that can be seen in the QSM images. With the utilization of the QSM term during the matching pursuit process, these strong negative values translate into high MVF. The HCFM predicts strong magnetic field perturbations in the extracellular space if the fibers are approximately perpendicular to the main magnetic field (red and green fibers in the FA map). Therefore, the amount of decay in those areas can be explained by the existence of a moderate amount of myelin. In this way, orientation informed MIMM corrects for those overestimations in the fiber tracts oriented left-to-right or anterior-to-posterior. However, the fiber tracts oriented superior-to-inferior do not follow the HCFM prediction since they still manifest highly negative QSM values. This means that even though the field perturbation due to myelin is very small, the material susceptibility of the existing myelin sheaths is substantial as captured by QSM. A possible explanation of this situation can be the existence of fiber crossings in those areas that are not covered by the current model. This remains to be a limitation of this study that needs to be investigated in future studies.

Another limitation of this study is the utilization of the fixed T2 values for the myelin and intra-extracellular water pools. This was done to decrease the number of free variables to solve for and make the matching pursuit process more well-posed. Another limitation is that the fibers inside the voxels are assumed to have the same orientation. This assumption will break in voxels with fiber crossings. In future studies, the model can be improved by including multi orientation fibers and multi T2 water pools. The stability of the fitting can then be improved using spatial regularization.

## 6. Conclusion

A novel microstructure informed matching pursuit method called MIMM is developed to measure MVF based on detailed biophysical modeling of the human brain using HCFM and distinct susceptibility source inclusions. The MIMM method is superior to previously developed 3PCF based myelin quantification using mGRE signals in terms of accuracy, visual quality, and processing time. A significant linear correlation with the reference T2-relaxometry method is observed (r = 0.75, p < 0.0001) whereas a significant correlation is observed with the 3PCF with lower correlation (r = 0.34, p < 0.0001). Furthermore, the feasibility of utilizing an atlas-based fiber orientation map was also investigated instead of a subject-specific fiber orientation map acquired using DTI since it may not be available in routine clinical scans. MIMM has the potential to be used as a myelin and iron quantification tool for longitudinal studies.

## Data statements

All images used in this work are de-identified to protect privacy of human participants. Code is available at https://github.com/MertSisman/MIMM. An example dataset is provided at https://zenodo.org/record/8193673. Remaining data is available to interested researchers upon reasonable request.

## CRediT authorship contribution statement

**Mert Şişman:** Conceptualization, Methodology, Software, Validation, Formal analysis, Investigation, Data curation, Writing - original draft, Visualization. **Thanh D. Nguyen:** Conceptualization, Methodology, Resources, Data Curation, Writing - Review & Editing, Supervision. **Alexandra G. Roberts:** Software. **Dominick J. Romano:** Conceptualization. **Alexey V. Dimov:** Conceptualization, Supervision. **Ilhami Kovanlikaya:** Conceptualization, Supervision. **Pascal Spincemaille:** Conceptualization, Methodology, Resources, Writing - Review & Editing, Supervision. **Yi Wang:** Conceptualization, Methodology, Writing - review & editing, Supervision, Funding acquisition.

## Declaration of Competing Interest

A.V.D., P.S. and Y.W. are inventors of QSM-related patents issued to Cornell University. P.S. and Y.W. hold equity in Medimagemetric LLC. T.D.N. and P.S. are paid consultants for Medimagemetric LLC.

## Acknowledgments

This work was supported in part by research grants from the NIH: R01NS105144, R01NS090464, R01NS095562, S10OD021782, R01HL151686, and National MS Society: RG-1602-07671.

## SUPPLEMENTARY MATERIALS

**Figure S1.**
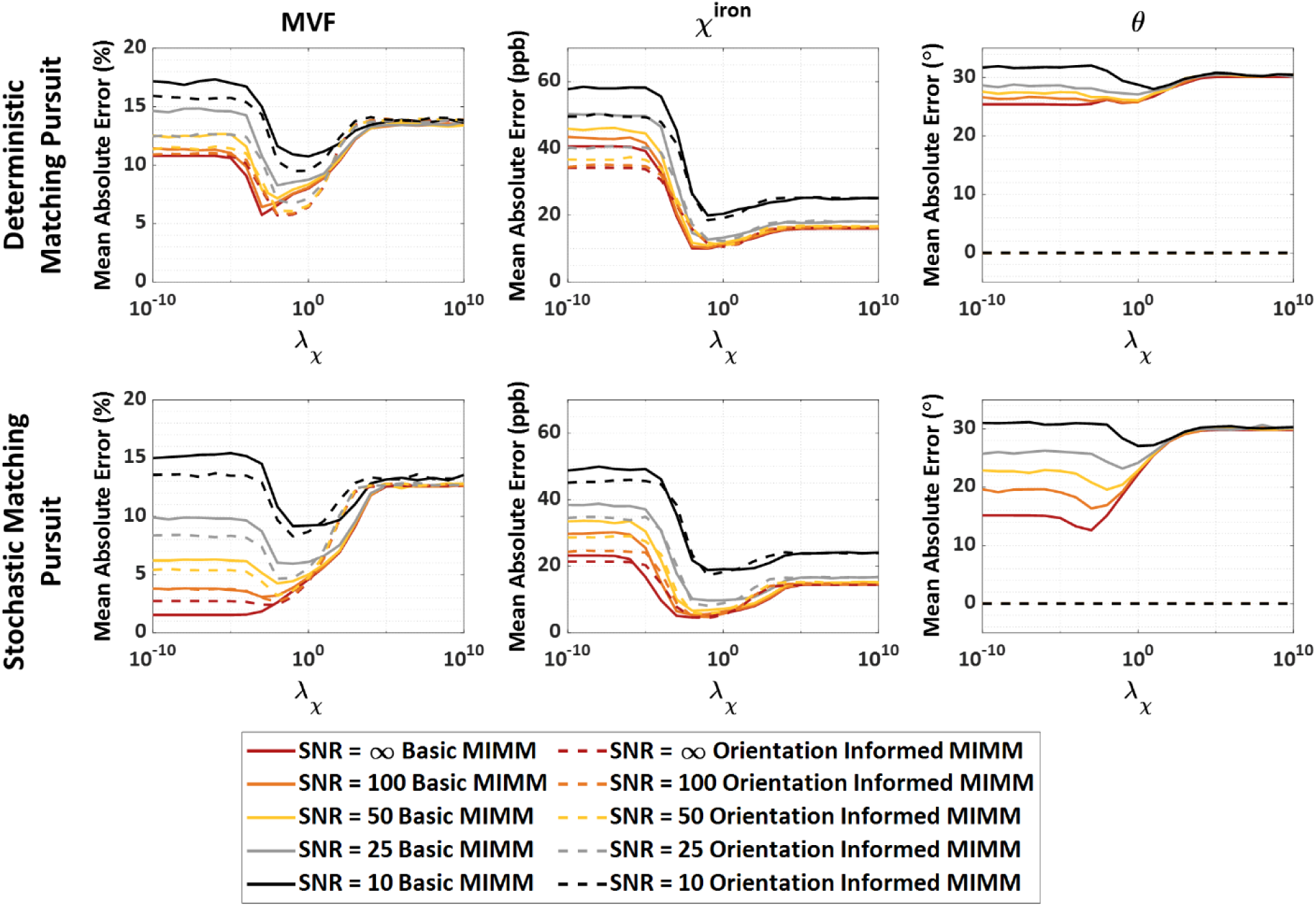
Monte Carlo simulation results using both the deterministic (top row) and stochastic (bottom row) matching pursuit to demonstrate the SNR performance of the algorithms with different *λ*_*χ*_ values. Columns show the mean absolute errors for the 3 outputs of the matching pursuit process: MVF, *χ*^*iron*^, and *θ*. Solid lines show the errors for the basic matching pursuit whereas dashed lines show the errors for orientation informed matching pursuit for 5 different SNR levels.

**Figure S2.**
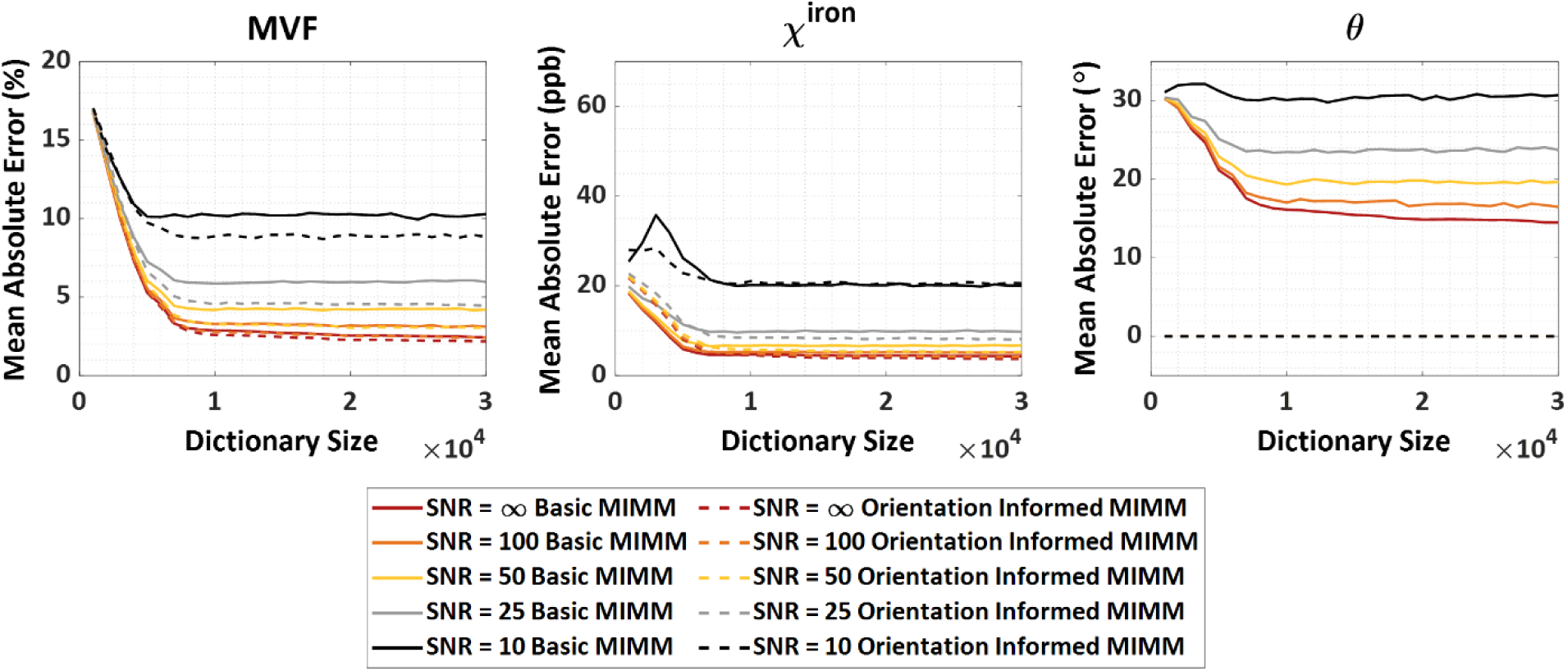
Monte Carlo simulation results to demonstrate the SNR performance of MIMM with different dictionary sizes. Columns show the mean absolute errors for the 3 outputs of the matching pursuit process: MVF, *χ*^*iron*^, and *θ*. Solid lines show the errors for the basic MIMM whereas dashed lines show the errors for orientation informed MIMM for 5 different SNR levels.

**Figure S3.**
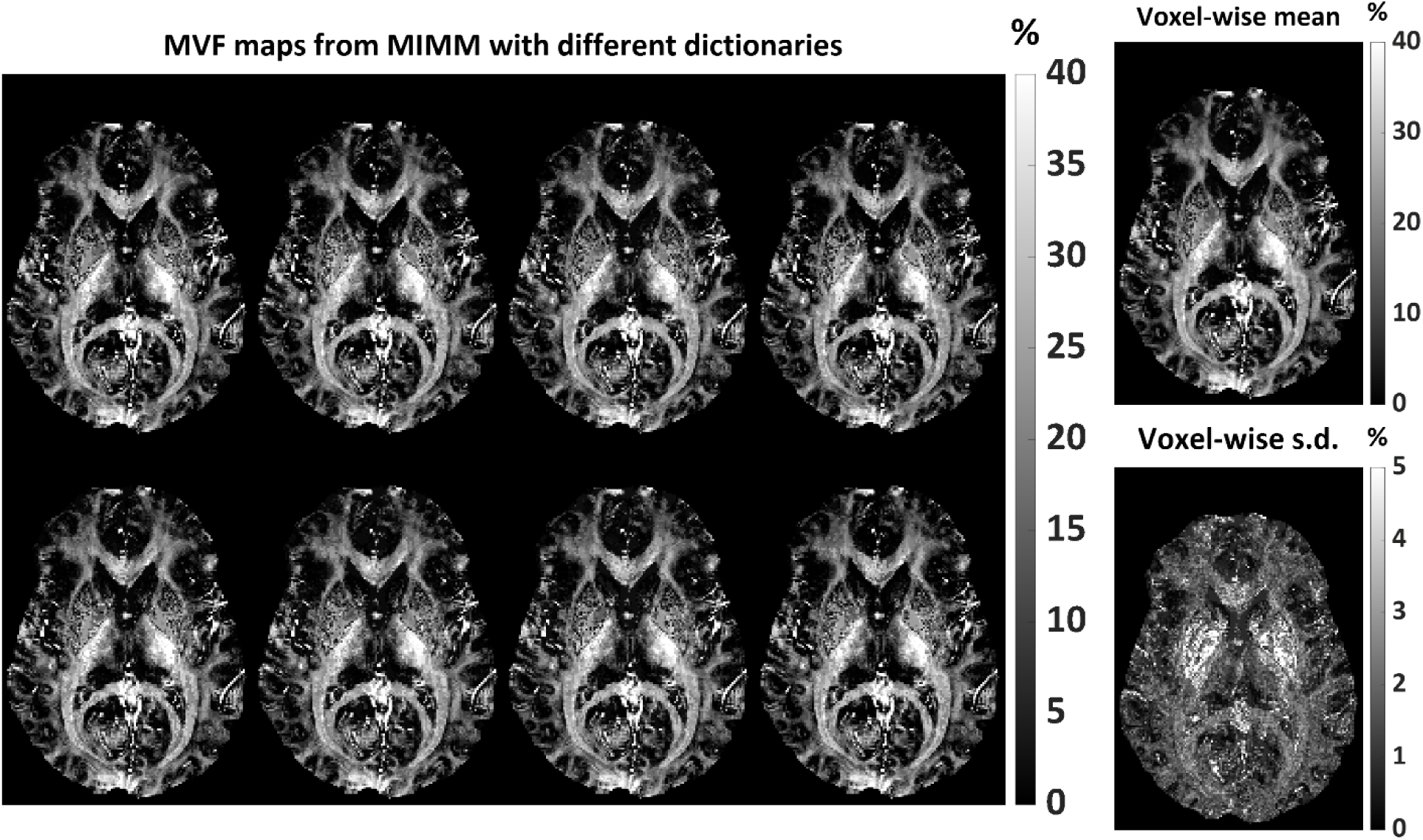
MVF maps obtained using orientation informed MIMM using DTI and 8 different and independently created dictionaries of size 10000 (left). *λ*_*χ*_ = 0.015 is used. The voxel-wise mean and standard deviation (S.D.) maps of the 8 MVF maps (right).

**Figure S4.**
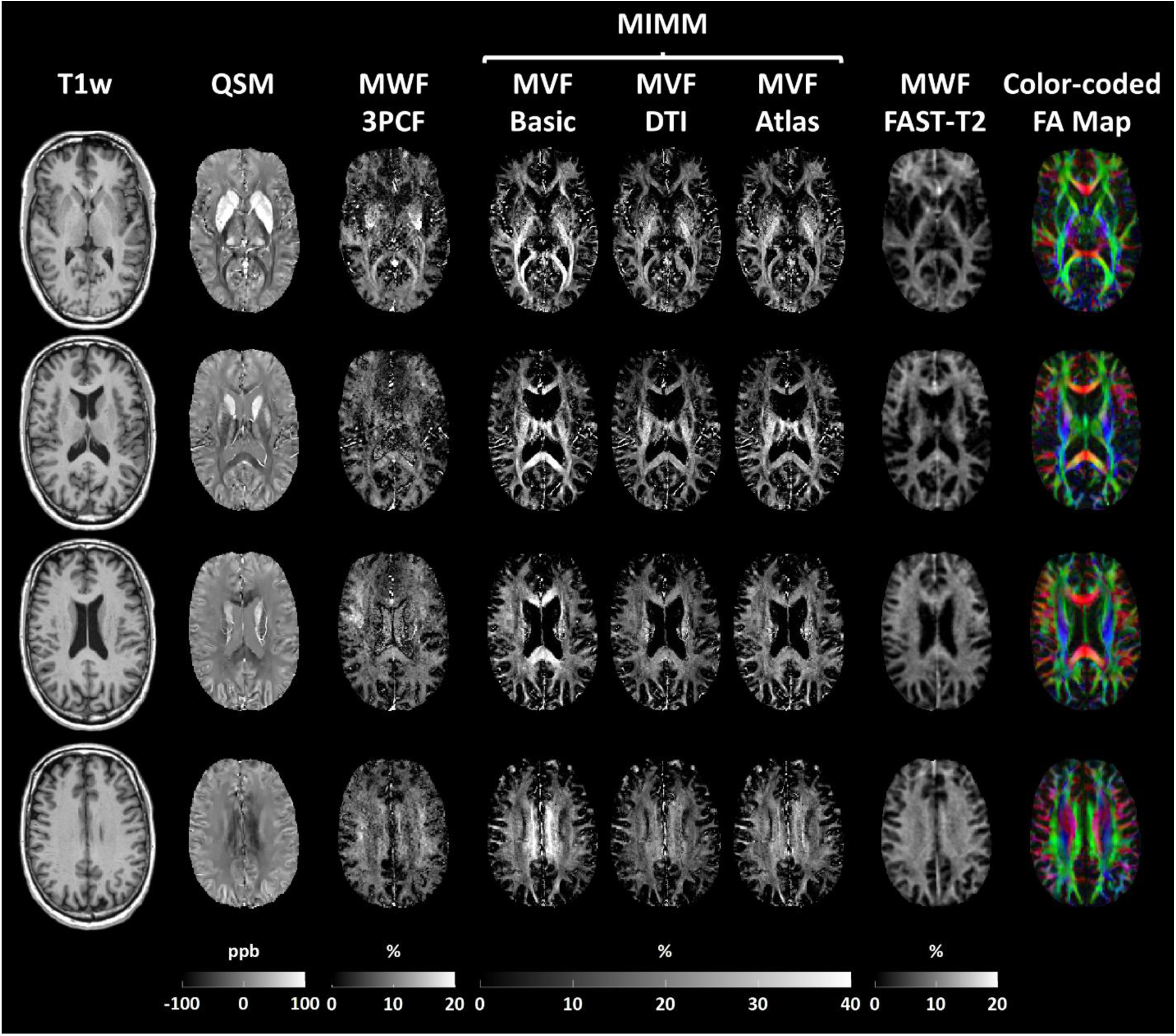
Example results from another subject. From left to right: T1-weighted images (T1w), QSM maps, MWF maps obtained with 3 pool complex fitting (3PCF), MVF maps obtained with basic MIMM, DTI orientation informed MIMM, and atlas orientation informed MIMM, MWF maps obtained using T2-relaxometry from the FAST-T2 data, and FA maps obtained from diffusion tensor images. All images are registered to the QSM space.

**Figure S5.**
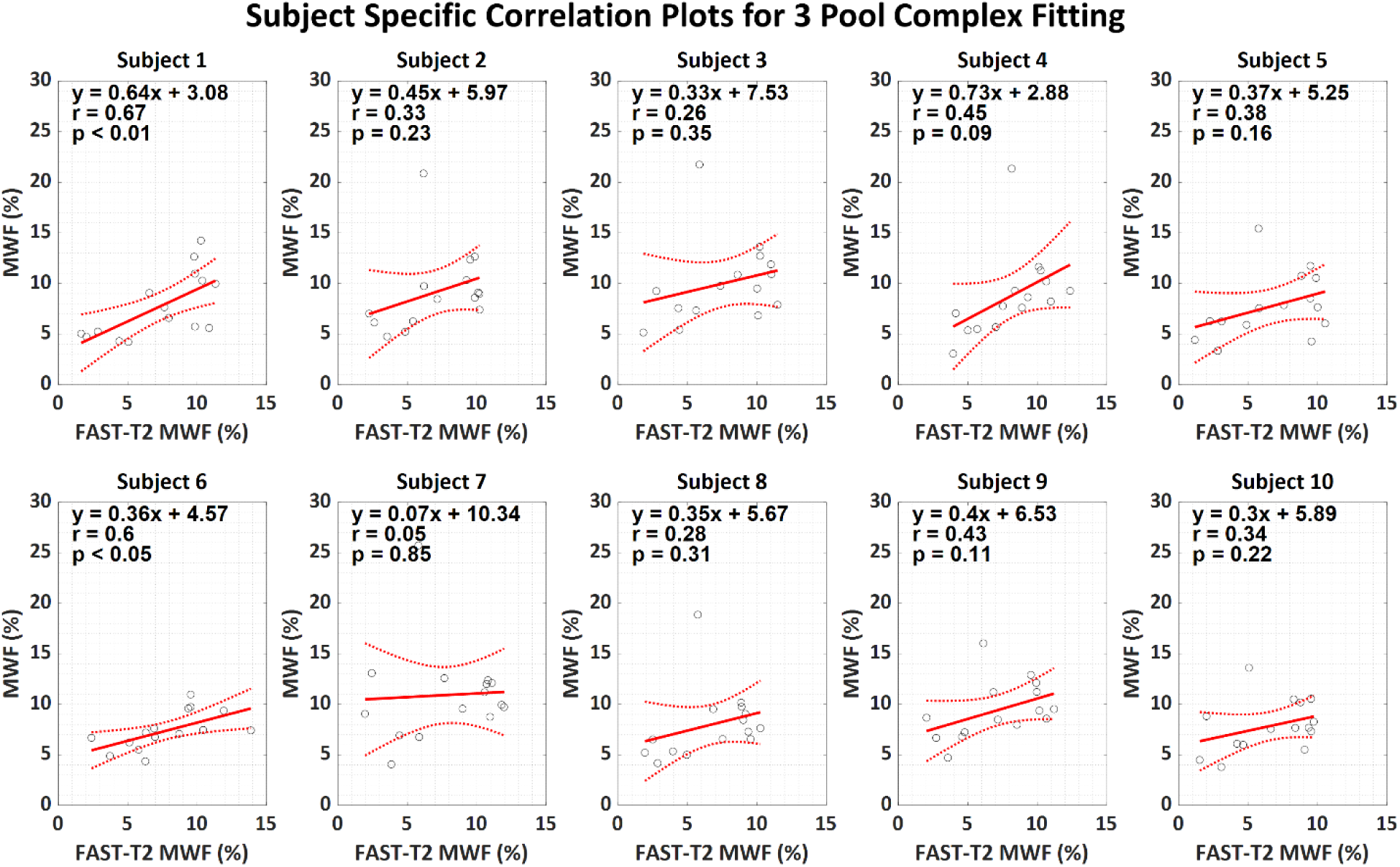
Subject-specific linear correlation analysis results between FAST-T2 based MWF maps and MWF maps obtained with 3 pool complex fitting.

**Figure S6.**
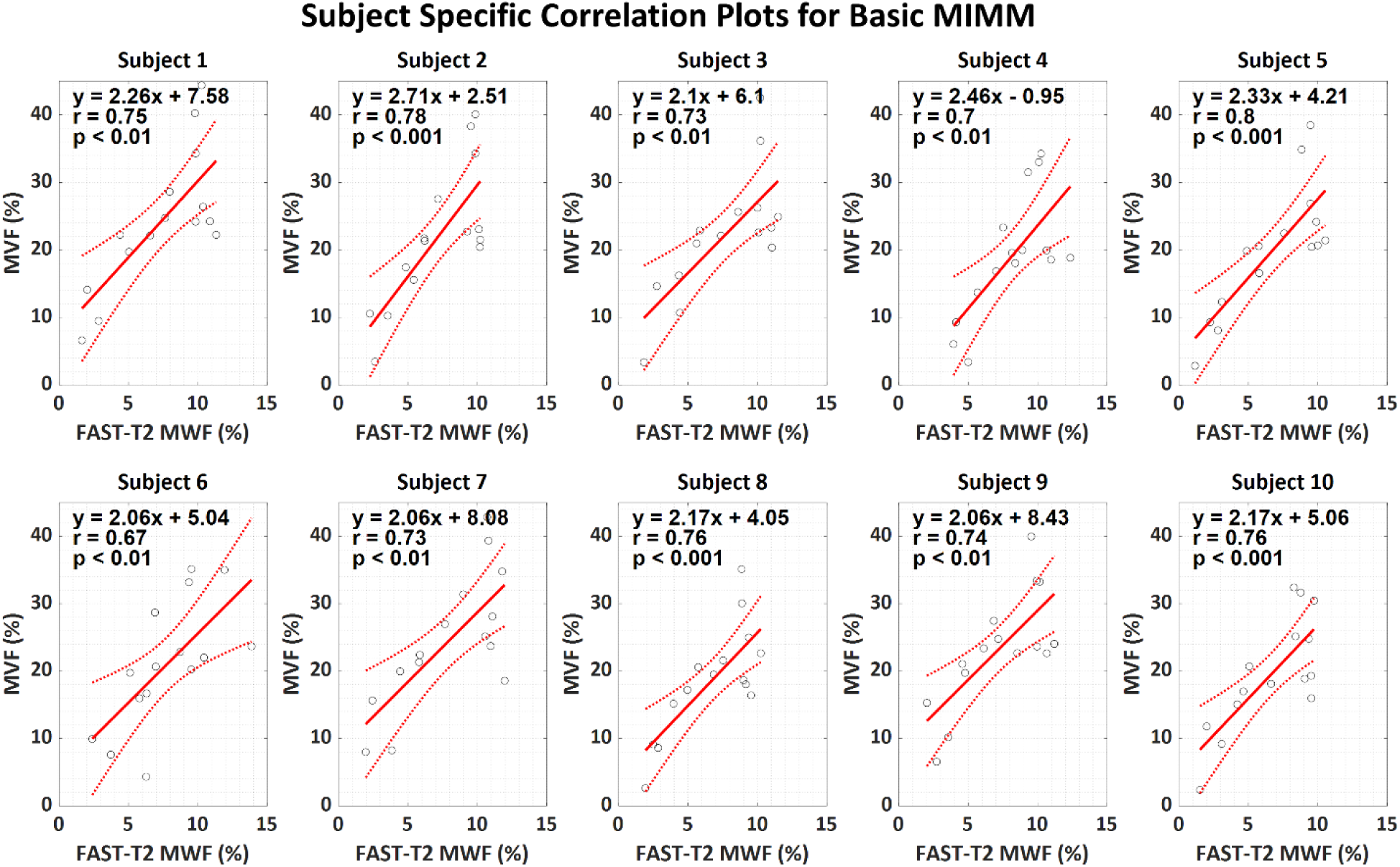
Subject-specific linear correlation analysis results between FAST-T2 based MWF maps and MVF maps obtained with basic MIMM.

**Figure S7.**
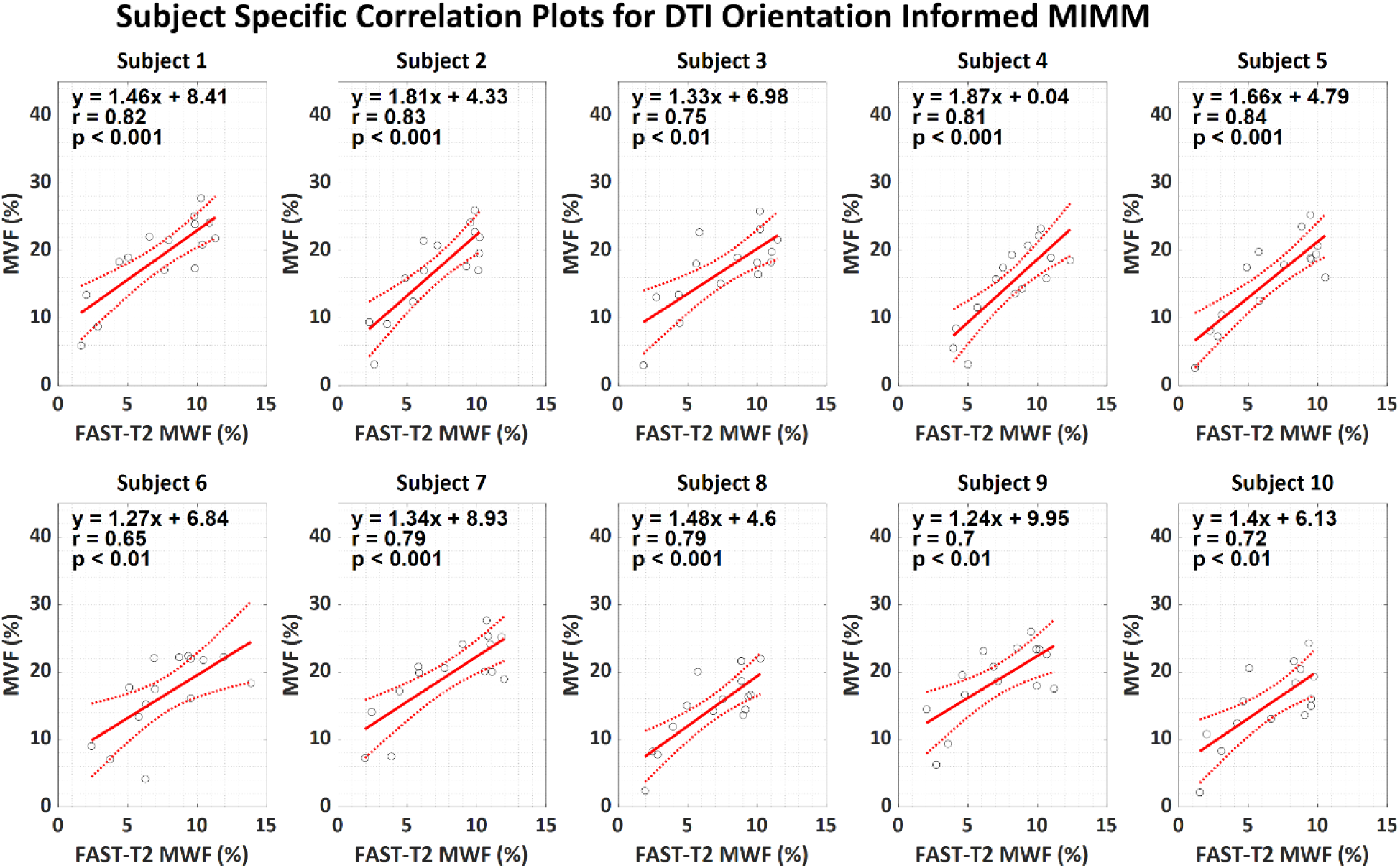
Subject-specific linear correlation analysis results between FAST-T2 based MWF maps and MVF maps obtained with DTI orientation informed MIMM.

**Figure S8.**
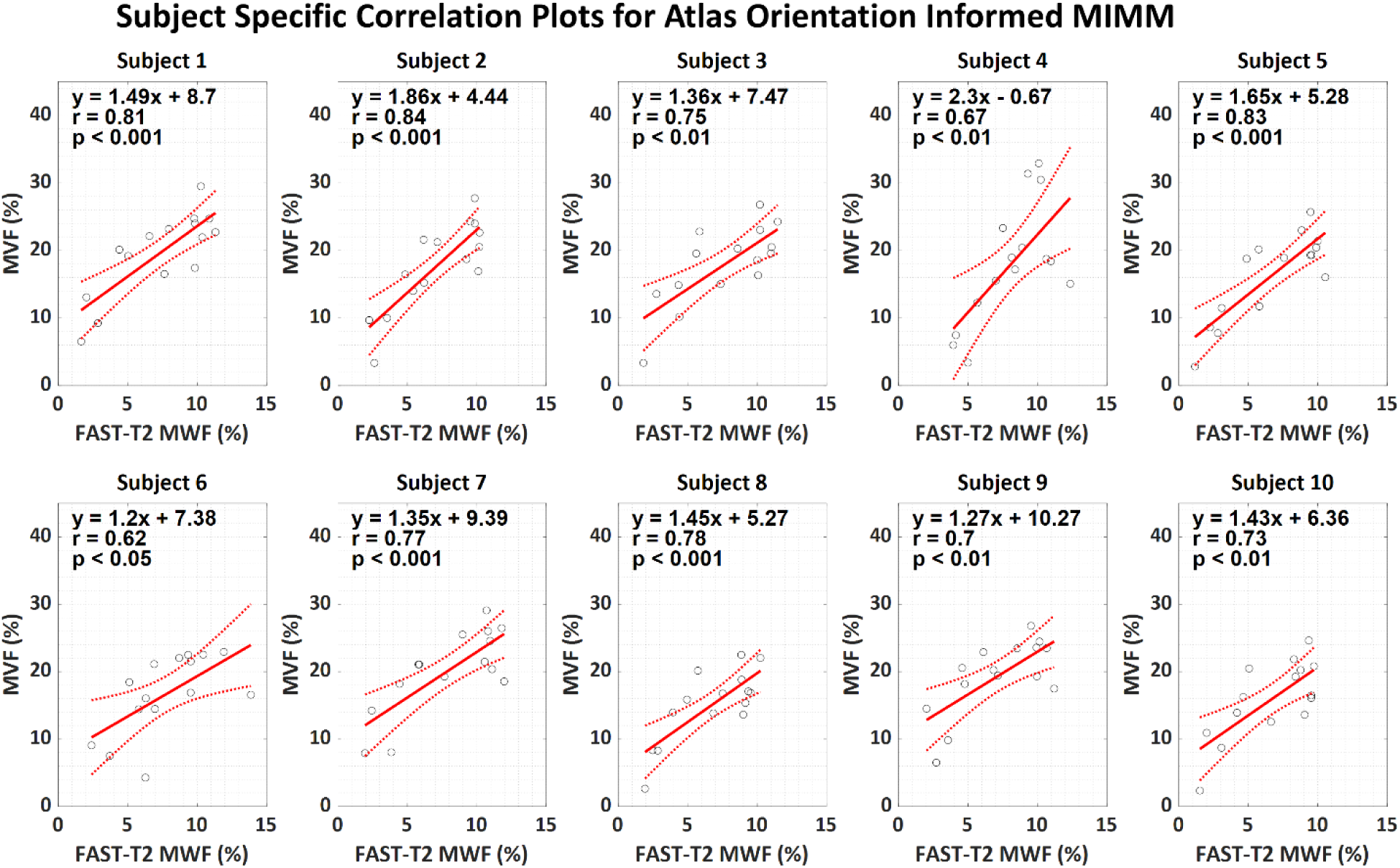
Subject-specific linear correlation analysis results between FAST-T2 based MWF maps and MVF maps obtained with atlas orientation informed MIMM.

## Notes

### Author Declarations

IRB of Weill Cornell Medicine gave ethical approval for this work.

